# Race/ethnicity differences in vitamin D levels and impact on cardiovascular disease, bone health, and oral health

**DOI:** 10.1101/2021.01.02.21249149

**Authors:** Ajoy Thamattoor

## Abstract

Vitamin D and its biomarker 25(OH)D are known to vary by race/ethnicity with African Americans (AAs) having significantly lower levels than non-Hispanic whites (white Americans). However, AAs have better bone mineral density (BMD) and less arterial calcification, one marker of cardiovascular risk, than white Americans, with some studies showing higher vit. D levels harmful to AAs. This study analyzes NHANES data from 2011 to 2014, NHANES being a biennially published national survey of nearly 10,000 people, with interview, examination, and lab data components. The analyses, using count regression and linear regression models to avoid thresholding of variables, find that abdominal aortic calcification scores rise with 24(OH)D in white Americans, with no statistically significant effect in AAs; femoral BMD falls with 25(OH)D in both groups; osteoporotic fracture risks fall with 25(OH)D in white Americans; and periodontal attachment loss falls with rising 25(OH)D in both groups. Overall, higher 25(OH)D seems protective for oral and skeletal health in white Americans, protective for periodontal health in AAs, negative for their skeletal health, and negative for arterial calcification in white Americans, after controlling for the demographic factors of age and sex, the physiological elements of blood pressure and BMI, the biochemical variables of LDL and cholesterol levels, the socioeconomic indicators of income-to-poverty-level ratio and education levels, and the environmental influence of the season. As periodontitis is low on the disease hazard scale compared to arterial calcification and skeletal health, the results point to a lack of significant protection with rising OH(D) levels in AAs, even after their low base levels, and some harmful impact from those higher levels. That combination should trigger a closer look at the single population-wide vitamin D threshold of 30 to 50 ng/mL currently recommended in the US.

## INTRODUCTION

Vitamin D is a secosteroid hormone with wide-ranging impact in the human body. Originally, it was known to influence calcium homeostasis and hence bone formation, bone remodeling, and fracture risk. Later, it was found to affect dental and periodontal health, kidney function, the immune system, arteriosclerosis via arterial calcification, and even acute respiratory tract infections. Most of this is mediated via its role in the regulation of calcium homeostasis and via vitamin-D receptors (VDRs) present in many cells (Marquina et al., 2018).

Vitamin D is not technically a vitamin as the body manufactures it endogenously from 7-dehydrocholesterol via the effect of sunlight on skin, an evolutionarily ancient mechanism present in most land vertebrates (Holick and Chen, 2008). This intermediate, cholecalciferol or vitamin D-3, is then converted to 25-hydroxycholesterol, 25(OH)D, by the liver (Chang and Lee, 2019). In the blood, 25(OH)D circulates with 85 to 90% bound to vitamin-D-binding protein (VDBP), 10 to 15% loosely bound to albumin, and < .01% in free state (Yu et al., 2018). VDBP-bound 25(OH)D is likely not biologically available; however, total 25(OH)D is what most studies consider a measure of bioavailable vitamin D. The proximal tubules of the kidneys convert this to 1,25-dihydroxycholesterol, also called 1,25(OH)2D3, the bioactive form. VDRs bind 1,25-dihydroxycholesterol (Chang and Lee, 2019).

The cutaneous synthesis pathway depends on the UV-B spectrum in sunlight. This pathway is less efficient on darker skin and in northern latitudes with less UV-B. African Americans (AAs), in particular, show statistically lower levels of 25(OH)D than white Americans; the mean serum level in one study was 15.6 ± 0.2 ng/mL as opposed to 25.8 ± 0.4 ng/mL for white Americans. However, vitamin-D deficiency is defined without respect to race/ethnicity or skin color, with recommendations of either 20 ng/mL or 30 ng/mL (50 to 75 nmol/L), leading to a situation where AAs are predominantly considered vitamin-D deficient (Powe et al., 2013). Vitamin D reference ranges are set based on populations of European ancestry (Freedman and Register, 2012). Vitamin D deficiency in white Americans leads to higher fracture risk, worse arterial calcification, more dental and periodontal issues, and, less clearly, higher cancer and cardiovascular risk (Holick and Chen, 2008). With AAs the effects are more mixed and there is controversy over whether vitamin D supplementation harms or helps them (Brown et al., 2018). The deficiency recommendation is based, to some extent, on vit. D levels in sub-Saharan Africa being much higher than that of AAs, the mean all above 20 ng/mL and in Nigeria, Uganda, Malawi, Gabon, Zimbabwe, Botswana, the Ivory Coast, and Guinea-Bissau above 30 ng/mL (Mogire et al., 2020, *Fig. 2)*

**Figure 1.**
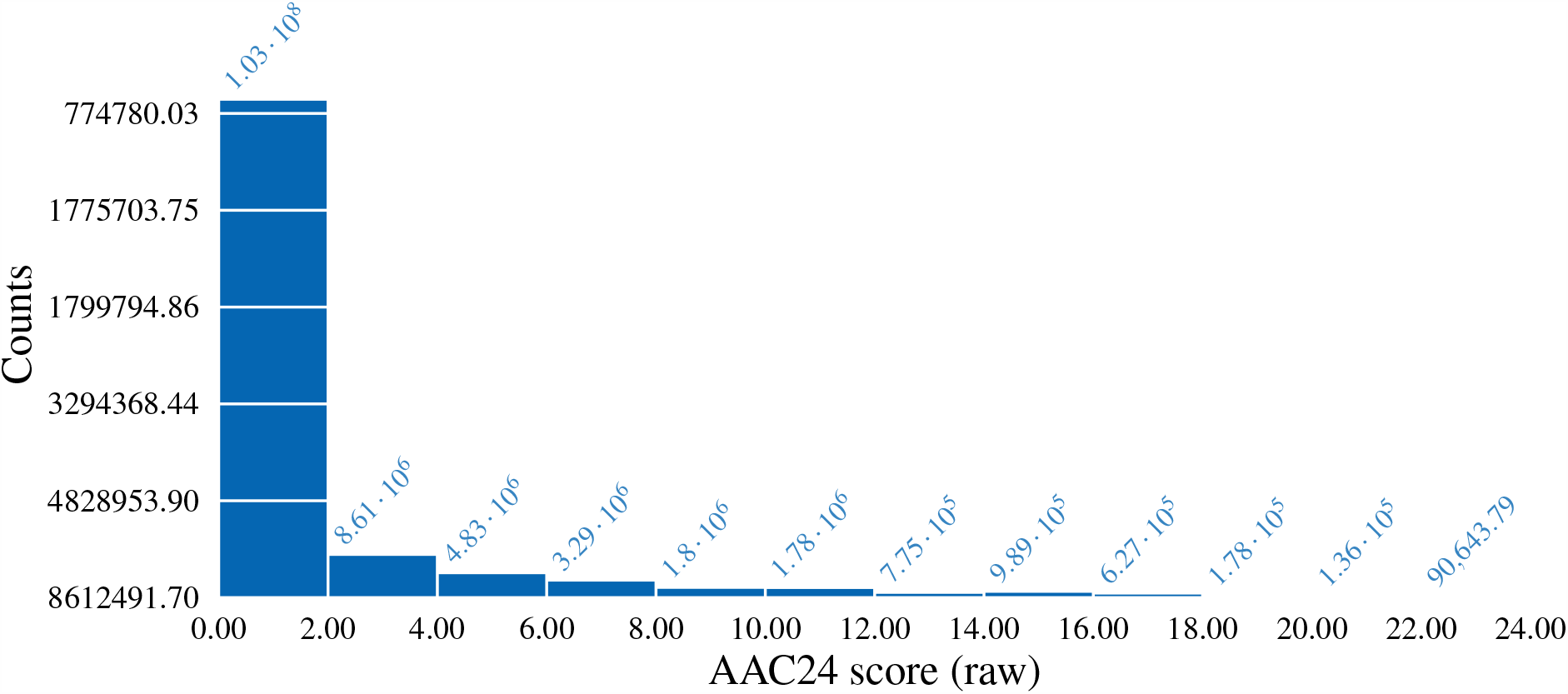
Histogram of AAC24 raw scores

**Figure 2.**
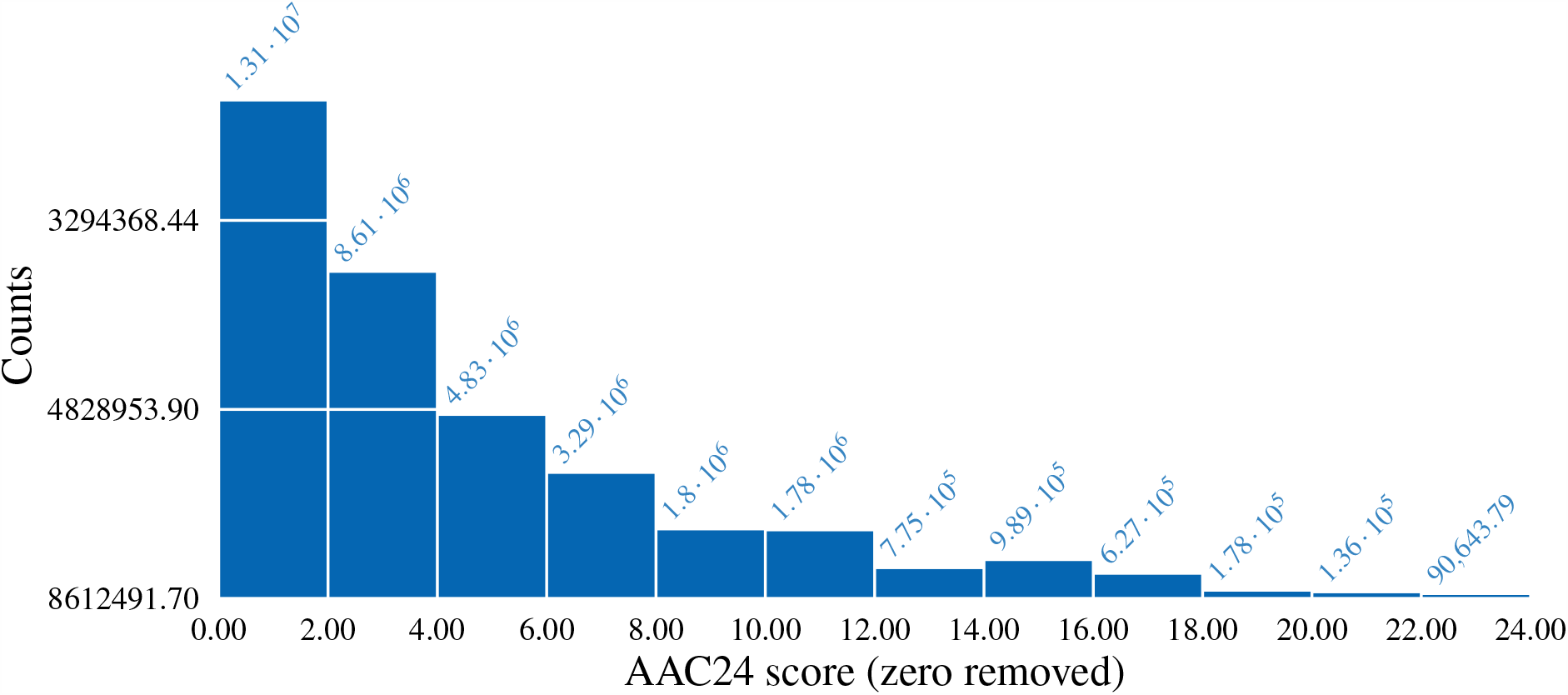
Histogram of AAC24 raw scores with 0 removed

Studies have shown that AAs have higher bone mineral density, more advantageous femur geometry in cortical bone and spongy trabeculae, lower risk of falls and fractures (Popp et al., 2017), and lower arterial calcification (though with higher cardiovascular disease, CVD, risk) than white Americans, despite having significantly lower 25(OH)D levels and, statistically, disadvantageous socioeconomic status (Newman et al., 2002; Freedman and Register, 2012). There is some evidence that higher 25(OH) levels increase CVD risk in AAs (Brown et al., 2018). For other effects of vitamin D deficiency, race/ethnicity differences are either not known or not seen. The protective effects in AAs may be from genetic causes such as ethnic-haplotype VDR polymorphisms more prevalent in them, VDBP gene polymorphisms, higher PTH levels (Powe et al., 2013), and genetic influences on trabecular microstructure and bone density (Popp et al., 2017).

One source of data to examine this discrepancy is the National Health and Nutrition Examination Survey (NHANES), conducted biennially by the National Center for Health Statistics (NCHS), an arm of the CDC. NHANES is a set of survey questions, physical exams, and laboratory tests on a nationally representative sample of around 5000 people annually. From 1998 to the 2013–14 cycle, NHANES included data on 25(OH)D serum levels; Orces et al. (2019) is one analysis of this data. NHANES also has data on periodontitis risk and prevalence for two of those cycles, 2011–12 and 2013–14; on abdominal aortic calcification and hip and osteoporotic fracture risk for 2013–14; hip and femur bone density for 2013–14; and whole-body bone mineral density (BMD) for 2011–12 and 2013–14. These data allow us to examine the role of vitamin D and race/ethnicity in these physiological measures and biochemical markers of health and predicted or already-present disease.

### Cardiovascular disease: Abdominal aortic calcification

Vitamin D is linked to CVD risk. But the results of studies looking for a link between vitamin D and markers of CVD and heart disease have been nuanced and mixed. A nationwide, randomized control study of vitamin D3 supplementation of 2000 IU per day, the VITAL study, showed no improvement in cardiovascular risk, with myocardial infarctions (MCIs), strokes, and death from CVD staying statistically the same across control and experimental groups even after a five-year follow-up. The results, or the lack of them, did not vary by race/ethnicity (Manson et al., 2019). NHANES-III data, collected from 1988 to 1994, showed an inverse association between 25(OH)D levels compared between highest and lowest quartiles and death from coronary heart disease (CHD), with no effect when all quartiles were considered. The effect remained after controls were added for sex, diabetes, income, age, BMI, blood pressure (BP), dietary elements, and similar lifestyle, socioeconomic, and physiological parameters (Daraghmeh et al., 2016).

A correlation between vitamin D levels and CVD may well indicate that low vitamin D is a marker of CVD than its cause. Vitamin D levels are affected by VDR gene polymorphisms, sex hormones and hence sex, age, use of corticosteroids, obesity, with fat sequestering the lipophilic vitamin, and kidney disease (Marquina et al., 2018). Thus, variations in the CYP24A1 and VDR genes have been associated with coronary artery calcification (CAC) (Shen et al., 2010); low 25(OH)D was correlated to heart failure rates in white but not AA adults in a study that spanned 20 years (Lutsey et al., 2015). Complementing that, Mendelian randomization studies have frequently failed to find a correlation between various SNPs in CYP genes and coronary artery disease, indicating vitamin D may be a biomarker instead of a causal element (Marquina et al., 2018).

Balanced against a null result for a relation between 25(OH)D and heart failure in AAs, the Jackson Heart Study, of 5000 AA adults in Jackson, Mississippi, showed a direct correlation of 25(OH)D and HDL cholesterol, no correlation with LDL, and inverse correlations with CVD risk factors such as BMI, waist circumference, BP, and fasting plasma glucose, all mediated partly through C-reactive protein and aldosterone. This is a pathway through which 25(OH)D can lower BP by downregulating the renin-angiotensin-aldosterone (RAAS) system by inhibiting renin synthesis, the start of the RAAS pathway. Lowering aldosterone reduces how much water and sodium the body retains, lowering blood pressure. The results of the path analysis held after controlling for age, sex, education, smoking, BMI, season, and other behavioral, socioeconomic and physiological elements (Khan et al., 2016). Large parts of these correlations were replicated in the overall population, including with no race/ethnicity control, with three cycles of NHANES data from 2001 to 2006. That study did not perform path analysis and so could not detect mediating influences (Ganji et al., 2020). Still another study with NHANES data from 2003 to 2006 found an inverse correlation between treatment-resistant hypertension, a major CVD risk, and 25(OH)D levels in the overall population (Alagacone et al., 2020). A third study found an inverse correlation between 25(OH)D levels and CVD mortality for the 1988 to 1994 NHANES sample, with participant mortality followed till 2000 (Melamed et al., 2008). Recent reviews have concluded that a J-shaped or U-shaped inverse correlation exists between CVD elements and 25(OH)D, that is, in some range there is a protective effect (Durup et al., 2015), but this relationship may well indicate unmodeled elements than any true biological effect (Grant et al., 2016). Vitamin D also regulates parathyroid hormone (PTH) as part of the calcium homeostasis loop involving renal tubules, the thyroid gland, and the parathyroid. Low levels of 25(OH)D lead to high levels of PTH; this can cause myocardial hypertrophy and secretion of more aldosterone, all raising CVD risk (Marquina et al., 2018).

The effects of vitamin D are, however, complex. As noted before, randomized control trials have largely failed to show CVD benefits from vitamin D supplementation (Zitterman, 2018). A recent meta analysis of vitamin D supplementation and CVD risk across 21 randomized clinical trials found that supplementation had no benefits with regards to cardiovascular events and related deaths, strokes, MCI, or even overall mortality (Barbarawi et al., 2019). A case-cohort study of 79,705 postmenopausal females in the multi-ethnic Women’s Health Initiative Observational Study (WHI-OS) found no correlation between CVD risk and total 25(OH)D, free 25(OH)D, or bioavailable 25(OH)D in either white or AA females. The researchers found no PTH link to CVD risk in AAs (though they did find one in white females) (Zhang et al., 2019). The 6436-participant Multi Ethnic Study of Atherosclerosis (MESA), which weighted ethnic/racial minorities up in its sample, found an inverse link between 25(OH)D and CHD event applying to white adults but not AAs. The authors speculate that the cause may be an interruption of the RAAS sequence that leads to more aldosterone with lower 25(OH)D. AAs have higher circulating 1,25-dihydroxyvitamin D, likely because of increased activity by the hydroxylase coded for by the CYP27B1 gene, polymorphic partly by ethnicity. On the catabolism side, the hydroxylase responsible has reduced activity in AAs. VDR binding strength too may differ by race/ethnicity, with some genes here polymorphic by race/ethnicity. In addition, AAs have statistically higher rates of PTH, added to their lower levels of 25(OH)D (Robinson-Cohen et al., 2013). Nevertheless, they have lower levels of arterial calcification, particularly CAC, one CVD risk element. This is despite their having higher scores on CVD risk factors such as BP, diabetes, and smoking rates (see Table 1 of Orimoloye et al. (2018)).

**Table 1.** AAC vs race, vit D, smoking, income, edu., BMI, LDL, glucose, sys. BP, age, sex

|  | Model 1 |
| --- | --- |
| (Intercept):1 | −1.664 (0.019)*** |
| (Intercept):2 | 0.781 (0.052)*** |
| (Intercept):3 | −0.069 (0.016)*** |
| RIDRETH1 | −0.025 (0.085) |
| blood_pressure_systole | 0.000 (0.001) |
| LBDLDL | −0.001 (0.000)*** |
| DMDEDUC2 | −0.108 (0.020)*** |
| LBXVIDMS | 0.002 (0.000)*** |
| RIDEXMON | −0.185 (0.034)*** |
| BMXBMI | −0.021 (0.006)*** |
| cig_smoking | 0.001 (0.000)*** |
| LBXGLU | 0.002 (0.001)** |
| INDFMPIR | 0.013 (0.019) |
| RIDAGEYR | 0.059 (0.001)*** |
| RIAGENDR | 0.053 (0.038)* |
| AIC | 140387545.518 |
| Log likelihood | −70193757.759 |
| Num. obs. | 444.000 |

\*\*\* $p < .001$ ; \*\* $p < .01$ ; \* $p < .05$

For measuring CVD risk, CAC has proved a durable indicator. The standard measure for CVD risk, the Framingham Risk Score (FRS), which incorporates sex, age, BP, total cholesterol, LDL, HDL, smoking status, and diabetes status, predicts CVD moderately well for white and AA subjects (D’Agostino et al., 2001). The coronary calcium score, obtained with CT scans, provides even more predictive information for CVD events and CHD for both white Americans and AAs, both males and females. This despite the fact that the incidence and the score itself differ statistically between race/ethnicities, but within a race/ethnicity it still provides a good CVD-prediction marker when added to the FRS (Detrano et al., 2008). It also allows the researcher/clinician to steer clear of self-report data, with its biases and inaccuracies. Another study with the MESA cohort has concluded that what matters for CVD risk is the absolute CAC score within a group and not relative amounts for the individual compared to the group as a whole (Budoff et al., 2009). Calcified atherosclerotic plaque, in general, has been linked to higher 25(OH)D and 1,25(OH)D levels in AAs, with overall levels of atherosclerotic plaque much lower in them than in white population samples (Freedman and Register, 2012). A study of a MESA subsample showed that AAs with greater share of European ancestry had higher prevalence and amounts of CAC (though with lower scores on carotid intima-media thickness, another marker of CVD), indicating a genetic link (Wassel et al., 2009).

Arterial calcification in the abdominal aorta (AAC) works similar to CAC in predicting cardiovascular risk. AAC predicts both CVD risk and fracture risk, providing a mechanistic explanation for a noted link between CVD and bone fragility. The link is particularly strong between hip and vertebral fractures and MCI, stroke, and heart failure (Szulc, 2016). AAC has been linked to 25-OH(D) deficiency, albeit mediated by age, in a sample of 769 Asian females, 75% of whom were menopausal. This study confirmed an inverse association between AAC and bone loss (Kim et al., 2012). Another study in Rabat, Morocco failed to find a link between vitamin D status and AAC in 429 postmenopausal females. However, this study used threshold values to categorize participants as hypovitaminosis, insufficient, and sufficient categories, with 10 ng/mL and 20 ng/mL as thresholds, and then used chi-squared tests and ANOVA to check the means (El Maghraoui et al., 2018). In AA specifically, vitamin D supplementation was found to have no influence on AAC in females over 60, in the PODA trial (Brahmbhatt et al., 2020). However, in a study in Tartu, Estonia, of 152 males over the age of 60, 25(OH)D levels were found to be correlated to aortic calcification, with the entire aorta (thoracic, abdominal, and pelvic) checked up to the bifurcation into the iliac arteries (Zagura et al., 2011).

The literature and the data thus lead to a hypothesis filling a gap testable with NHANES 2011–14 data. *Hypothesis 1: AAC is directly linked to 25(OH)D levels in white Americans but unlinked in AAs*.

### Bone health: Bone mineral density and fracture risks

Bone fracture risk has many components. A risk score, the FRAX score, based on cohort studies in multiple countries and ethnicities, was developed by the University of Sheffield, UK, in 2008, based on data collected in collaboration with the World Health Organization. This tool uses an algorithm to weight risk factors using t-scores and z-scores, that is, deviations from an average, to come up with a risk number. For the US, the risk elements for people not taking any medication include age, sex, BMI, individual and family history of fracture, smoking status, three or more alcoholic drinks a day, rheumatoid arthritis, and areal BMD (aBMD). The tool weights the risk differently for races/ethnicities (Center for Metabolic Bone Diseases, 2008). FRAX predicts hip fractures well but not necessarily other types of fractures, because some such as spine fractures do not get even reported or diagnosed. Biochemical markers such as 25OH(D) are not included; all the measures included are detected noninvasively (Unnanuntana et al., 2010).

The effect of vitamin D on bone is from its role in calcium balance. When serum CA^2+^ levels are low, the parathyroid gland detects this and increases production of PTH. PTH, in turn, stimulates renal CYP27B1, which hydroxylates 25(OH)D to 1,25(OH)D, which, in turn, inhibits both CYP27B1 and PTH in a negative feedback loop. In another feedback loop, 1,25(OH)D stimulates FGF23 in bone, which too inhibits CYP27B1. The catabolism of 1,25(OH)D, which usually exists for just a few hours, is triggered by the dehydroxylase CYP24A1. This dehydroxylase exists in all cells with VDRs (a nuclear receptor), and is stimulated by the presence of 1,25(OH)D. In the kidneys alone, CYP24A1 is inhibited by low CA^2+^ and PTH, that is, inversely with CYP27B1, the anabolic hydroxylase (Christakos et al., 2020).

Vitamin D facilitates CA^2+^ absorption in the intestine and, along with PTH, in the distal renal tubules and collecting tubules. Absorption is via the CA^2+^ channel TRPV5 and calbindin and excretion via CA^2+^ ATPases and Na^+^/CA^2+^ antiporters. VDRs are also expressed in osteoblasts and osteocytes but to a lesser extent in osteoclasts and chondrocytes. When serum CA^2+^ levels fall, both PTH and 1,25(OH)D levels rise, and this leads to bone resorption to replenish CA^2+^ levels. The resorption is mediated by expression of RANKL, which stimulates osteoclast differentiation and transfer of CA^2+^ from cortical and trabecular bone to serum. In parallel, bone remineralization is inhibited by the rise in pyrophosphate levels induced by 1,25(OH)D (Christakos et al., 2020). Most 25(OH)D in the blood is bound to VDBP; it is the free 25(OH)D that binds to VDRs and affects bone remodeling (LeBoff et al., 2020).

Bone mineral density, specifically aBMD, the area-wise density measured by DEXA, is strongly linked to fracture risk. Some studies indicate volumetric BMD too is linked to fracture risk, but this element is harder to measure, requiring quantitative computerized tomography (qCT). A recent meta-analysis of aBMD and fractures in studies looking at patients undergoing osteoporosis treatments showed a strong association of hip, femoral-neck, and lumbar-spine BMD with vertebral fractures and a less strong but significant association with hip fractures. For nonvertebral fractures, the heterogeneity of the definitions of such fractures and the occurrence of trauma fractures made the analysis not useful (Bouxsein et al., 2019). BMD itself depends, among other elements, on 25(OH)D levels: An analysis of NHANES III data found BMD to rise with 25(OH)D in all races/ethnicities (Bischoff-Ferrari et al., 2004). Even though BMD and Vitamin-D are linked, vitamin D supplementation, has had no effect on either aBMD measured with DEXA scans or vBMD and cortical and trabecular microstructure measured with peripheral quantitative CT scans. These null results, from the VITAL study among others, have held across age, race/ethnicity, and sex (LeBoff et al., 2020). Even higher doses of vitamin D, 4000 IU/day, led to reduced vBMD in the radius bone, and still higher doses in the tibia as well. This, however, must be considered more an instance of vitamin D poisoning from overdose (Burt et al., 2019).

Vitamin D interacts with race/ethnicity in its effect on BMD. AAs have higher BMD and lower risk of osteoporosis than white Americans though AAs have statistically lower levels of 25(OH)D. Though environmental and physical elements such as higher levels of obesity play a role in the lower 25(OH)D levels in AAs, as adipose tissue can sequester vitamin D or volumetrically dilute it, a large part is considered to be genetic. AAs also have statistically lower consumption of calcium, both from social elements and from the absence of the lactase enzyme making it harder to consume lactose-containing milk (Freedman and Register, 2012). Despite that, BMD as measured by DEXA, which measures largely aBMD, and vBMD measured by qCT, are higher in AAs than in others (Bachrach et al., 2019). QCT has shown higher cancellous, that is, trabecular, bone density in the spine in AA children than in white children; at the femoral shaft, AA children show greater bone area but no difference in cortical thickness. The active biochemical vitamin D marker measured in that study, 1,25(OH)D, showed a sex but no race/ethnicity difference and did not correlate with any bone measure (Gilsanz et al., 1998). Both larger bone mass and across-the-board volumetric density (vBMD) act as protective elements in AAs, lowering their osteoporosis and fracture risk.

The NHANES-III association between vitamin D and BMD was weaker than for the rest in AAs and found only among older adults. Its statistical significance vanished when physical activity was added to the list of confounders (Bischoff-Ferrari et al., 2004). This led the researchers to label BMD as the better endpoint marker for measuring bone health against 25(OH)D for non-AA populations, leaving the issue open for AA, especially older AA, populations (Bischoff-Ferrari et al., 2006). The higher BMD and lower 25(OH)D for AAs and the concomitant lower fracture risk does not bode well for a single 25(OH)D threshold for bone health across races/ethnicities, independent of whether the differences are environmental or inherited.

Gutiérrez et al. (2011) analyzed the relationship between vitamin D, PTH levels, and aBMD in NHANES data from 2003 to 2006. They found that BMD and 25(OH)D were directly related in non-AAs but not in AAs; PTH was inversely related to 25(OH)D in all groups but was maximally suppressed at a 25(OH)D level of 26 ng/mL in AAs. No measures of fracture risk were checked against either BMD or 25(OH)D.

These open issues and the availability of unanalyzed aBMD and FRAX data in NHANES 2011–12 and 2013–14 lead to the next hypothesis: *Levels of 25(OH)D are directly, that is, positively, related to spine and femur BMD and spine and hip FRAX in white populations, but no relationship exists in AAs in general*.

### Oral health: Periodontitis vulnerability

Periodontitis (PD) arises from loss of gum and supporting tissues for teeth, eventually leading to tooth loss. It is preceded by gingivitis—inflammation of the gum—and tissue loss follows from immune response to the oral microbiome, that is, the body’s response to microbial infection of the gums (Machado et al., 2020). PD is detected by probing of the pockets in gums and then adjusting that against recession of the gingiva to obtain a measure of attachment loss (AL), loss of tooth-supporting tissue. Thresholds are applied for a diagnosis of PD. The etiology of PD is multifactorial, with strong environmental and some genetic components. One strong environmental component is smoking. Studies have estimated 52% of PD in the US as due to current or former smoking, the 52% figure being an attributable fraction, that is, what proportion will not have PD if no one smoked. Almost 3*/*4^ths^ of PD risk in current smokers is from smoking, again considered as an attributable fraction (Tomar and Asma, 2000).

Meta studies have shown higher vitamin D reduces periodontitis, though the issue is still open. Vitamin D supplementation has unclear effect, again across studies. Vitamin D acts probably by binding to VDRs in gingival fibroblasts and periodontal ligament cells, inducing cathelicidin, an antimicrobial protein. This immune response is protective against PD (Machado et al., 2020). The association of cathelicidin and another antimicrobial, human beta defensin 2, with vitamin D is believed to be mediated by the binding of 1,25(OH)D to the VDR that then binds to the nuclear DNA sequence called the vitamin D response element (Wang et al., 2004). Vitamin D, however, has complex interactions with both the humoral and the cell-based immune systems, and these anti-inflammatory actions too might influence its effects on periodontitis (Dragonas et al., 2020).

An analysis of NHANES data from 1999 to 2004 estimated that higher levels of Vitamin D decreased the odds of periodontitis by 39% and 24% in former and non-smoker groups, respectively. Several demographic and environmental elements mediated the influence in the negative direction, that is, increasing the risk: AA race/ethnicity, older age, smoking status, male sex, and higher blood levels of retinyl stearate, retinyl palmitate, lead, non-coplanar polychlorinated biphenyls (PCBs) PCB105, PCB146, PCB172, PCB177, PCB178, PCB183, and PCB206, and higher urine levels of antimony. Higher levels of serum carotenoids and vitamin D had a protective influence (Emecen-Huja et al., 2019). Another analysis of NHANES III showed vitamin D and periodontitis correlated in both males and females over the age of 50, even after controlling for BMD, indicating mediation via the immunobeneficent role of vitamin D. This version of NHANES had just a half-mouth exam (random upper and lower quadrant) and only two sites per tooth. The researchers used vitamin D quintiles as the independent variable (IV) and raw mean AL as the dependent variable (DV), with a host of control variables (Dietrich et al., 2004).

Opposing this, the Dragonas et al. (2020) meta study mentions several studies showing a lack of effect for both 25(OH)D and 1,25(OH)D on periodontitis markers, as opposed to raw AL. However, this metastudy found a lack of result in studies largely from non-US populations, specifically from Finland; Korea; Pomerania, Germany; and Beijing, China. The one US study with a null result was on postmenopausal women. In some cases, the results were based on a linear regression against mean AL taken as a continuous variable. However, mean AL is not normally distributed and neither would be the measurement error, violating a requirement for ordinary least-squares linear regression. The environmental influences on periodontitis, and on vitamin D levels as well, are so significant that the heterogeneity in the meta review makes conclusions difficult. Another recent review by Millen and Pavlesen (2020) also found some support for a vitamin D role in periodontitis. This survey focused on cross-sectional studies, noting the paucity of longitudinal studies on the subject. Cross-sectional studies cannot establish the temporal direction of the connection; there are some indications gingivitis induces gum tissue to produce more 25(OH)D, which confounds both the overall expected result and the directionality of the influence.

Race/ethnicity differences in the occurrence of periodontitis are known. Adult AAs develop periodontitis at higher rates than white Americans do, with a large part of the causation likely environmental. An analysis of the 2009–10 NHANES data, which conducted full-mouth 6-site periodontal exams, showed higher rates, though not the highest rates, of periodontitis in AAs. Age, sex, smoking status, and socioeconomic-status elements such as education and income to federal poverty level (FPL) ratio were all associated with AL: older, smoking, less educated, and poorer males had higher AL (Eke et al., 2012). This study did not look at the influence of vitamin D on periodontitis; the earlier analysis of NHANES III by Dietrich et al. (2004) did look at that and found no differences across races/ethnicities, that is, stratification by race/ethnicity yielded results similar to that of the total. However, one notes that the half-mouth 3-site periodontal exam of NHANES III significantly underestimated overall periodontitis rates, assuming rates did not dramatically jump from 1994 to 2009 (Eke et al., 2012).

This leads to our final hypothesis: *Mean attachment loss is linked to vitamin D in both white and AA adult males and females, after controlling for age, sex, smoking status, education level, and family income*.

## METHODS

All analysis was done in R 4.3 using packages RNHANES for loading NHANES survey data, svyVGAM for running complex design-based GLM inference, survey for handling multi-stage sampled and weighted data such as NHANES, weights for weighted-variable histograms, xtable and texreg for getting R output directly into latex tables, and utility packages such as plyr, MASS, car, Hmisc, stringr, and stats. The coding environment was Rstudio 1.3 though there are no Rstudio dependencies in the methods or the code. The figures were generated using pgfplots, a latex package, and the tables as latex code directly output from R.

The central tool of our analyses is regression. The DVs vary: some are continuous, some discrete, some normally distributed, and some log-normal. The generalized linear model is appropriate. The exact regression model depends on the DV itself. If it is normally distributed, linear regression is appropriate. The requirement for linear regression is that the residuals be normally distributed, that is, *P* (*Y*| *X* = *x*) be normally distributed, where *Y* is the DV, *X* an IV, and *x* an arbitrary value of the IV. The conditional probability is unknown; we instead try to make *P* (*Y*) normally distributed, though in theory this is neither required nor sufficient, on the chance this makes *P* (*Y* |*X* = *x*) closer to normal. As we are interested largely in the presence of effects and less on their exact quantification, we use the multiplicative model, that is, log of original DV as the DV, where appropriate (Gustavsson et al., 2014). Normality applies, however, only to continuous distributions. If the DV is better modeled as discrete, then we are likely dealing with count data and must look at Poisson-distribution shapes and Poisson and similar regressions such as the negative binomial.

### DVs and models

For hypothesis 1, the DV is AAC24 in file DXXAAC, available for 2013–14. It has been found to be a reliable predictor for abdominal aortic calcification more accurately measured through other means, that is, via qCT scans. Note that AAC may contain both intimal and medial calcification, and medial calcification is not believed to be related to CVD risk. An advantage of AAC over qCT measures is that AAC is readily available as DEXA scans of the spine are recommended by several medical organizations to detect vertebral fractures in those older than 65 or 70 (DeSapri and Brook, 2020). AAC information is a byproduct of lateral scans of the spine. The variable itself is discrete with 24 values. It can be considered as count data, and its distribution does show a Poisson shape (see *Fig. 1*). The mean and variance are not equal, and hence a negative binomial distribution is a good model. The data have zero inflation, that is, bunching at the 0s beyond what the probability distribution would predict; hence the right GLM model to use is the zero-inflated negative binomial. *Figure 2* shows the shape of the data after zero removal, a Poisson/negative binomial shape. For svyVGAM package, that would be the svy vglm function with GLM function as zinegbinomialff. The design object for this call comes from the survey package’s svydesign() call with id being SDMVPSU, and strata being SDMVSTRA. The setting of weights, required in this function, is explained later.

For hypothesis 2, there exist four DVs: spine BMD DXXOSBMD in file DXXSPN, femur BMD DXXOFBMD in file DXXFEM, and hip and osteoporotic FRAX calculated from files DXXFRX. All are available for 2013–14 though only for those above the age of 40. Both spine and femur BMDs are roughly normal shaped (*Fig. 3, Fig. 4*) and the QQ plots are roughly lines (not shown), and so direct linear regression (svyglm call from survey) is used. This analysis eschews K-S and Shapiro Wilk tests for normality as these are mostly ineffective with large sample sizes. For the BMD variables, only aBMD information is available as this is a DEXA scan; vBMD and cortical and trabecular bone geometry require qCT scans. Hip FRAX is neither normal nor log-normal, so it was used as is (*Fig. 5*). Osteoporotic FRAX is Poisson-shaped but with unequal mean and variance and with no zero inflation (*Fig. 6*); hence survey vglm with negbinomial as the GLM is used. The hip FRAX is picked up from either DXXFRAX1 or DXXFRAX3 depending on the value in DXXPRVFX (prior fracture present or not). Osteoporotic FRAX is calculated similarly except that DXXFRAX2 or DXXFRAX4 is used.

**Figure 3.**
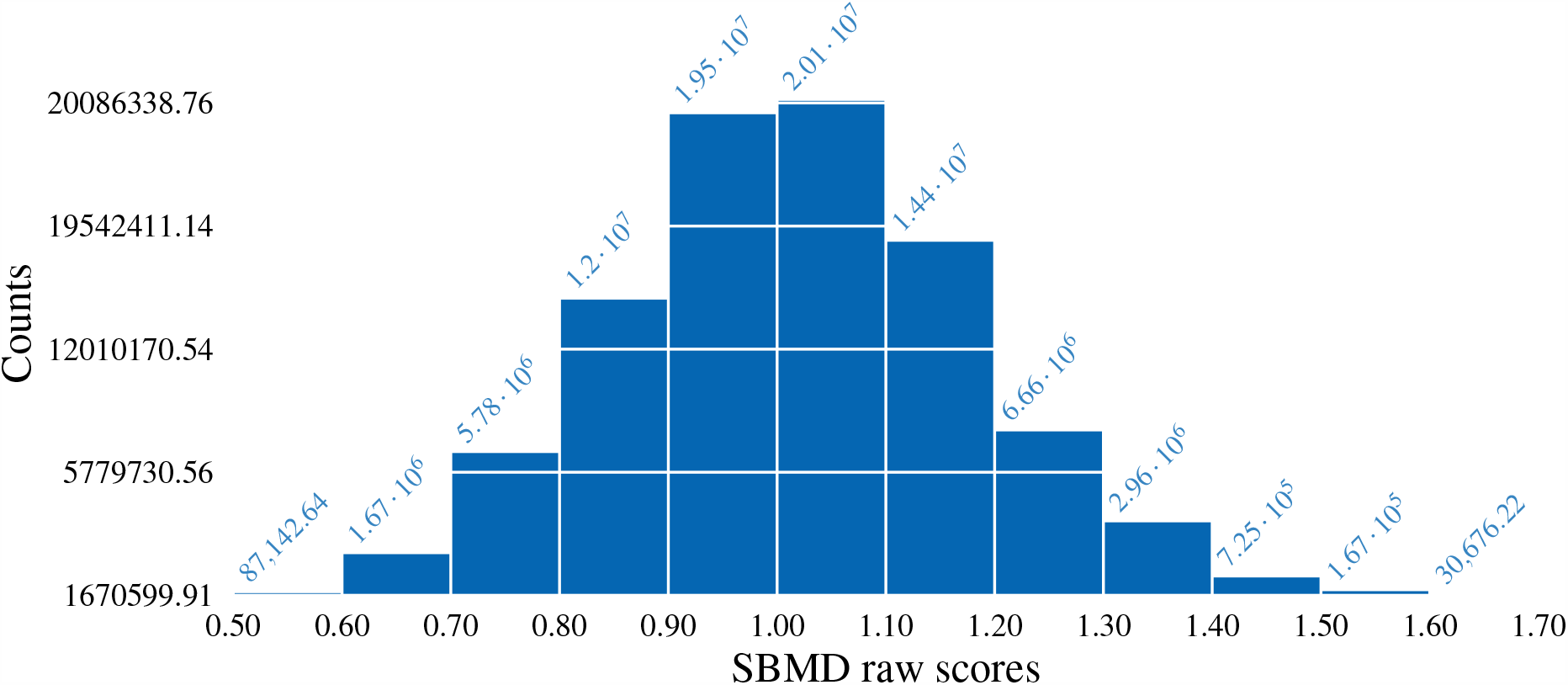
Histogram of spine-BMD raw scores

**Figure 4.**
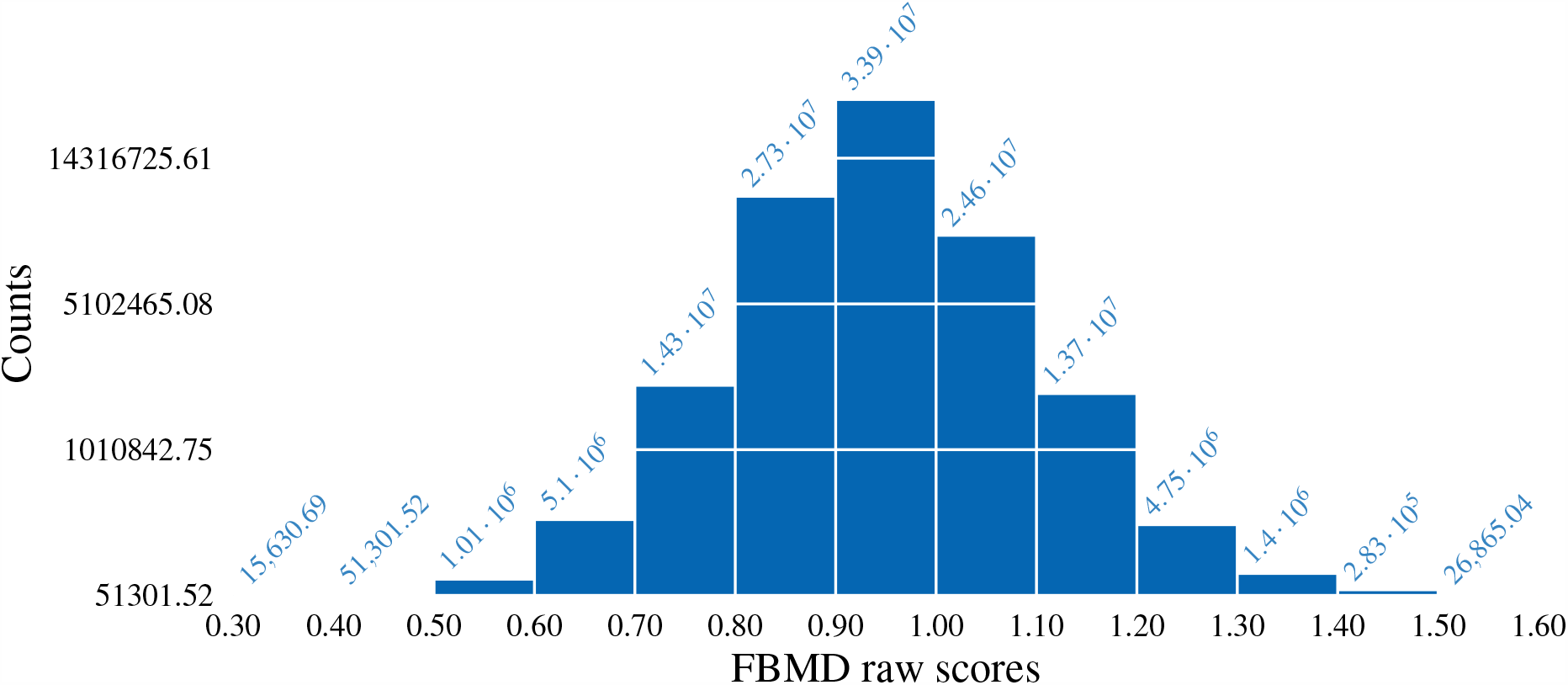
Histogram of femur-BMD raw scores

**Figure 5.**
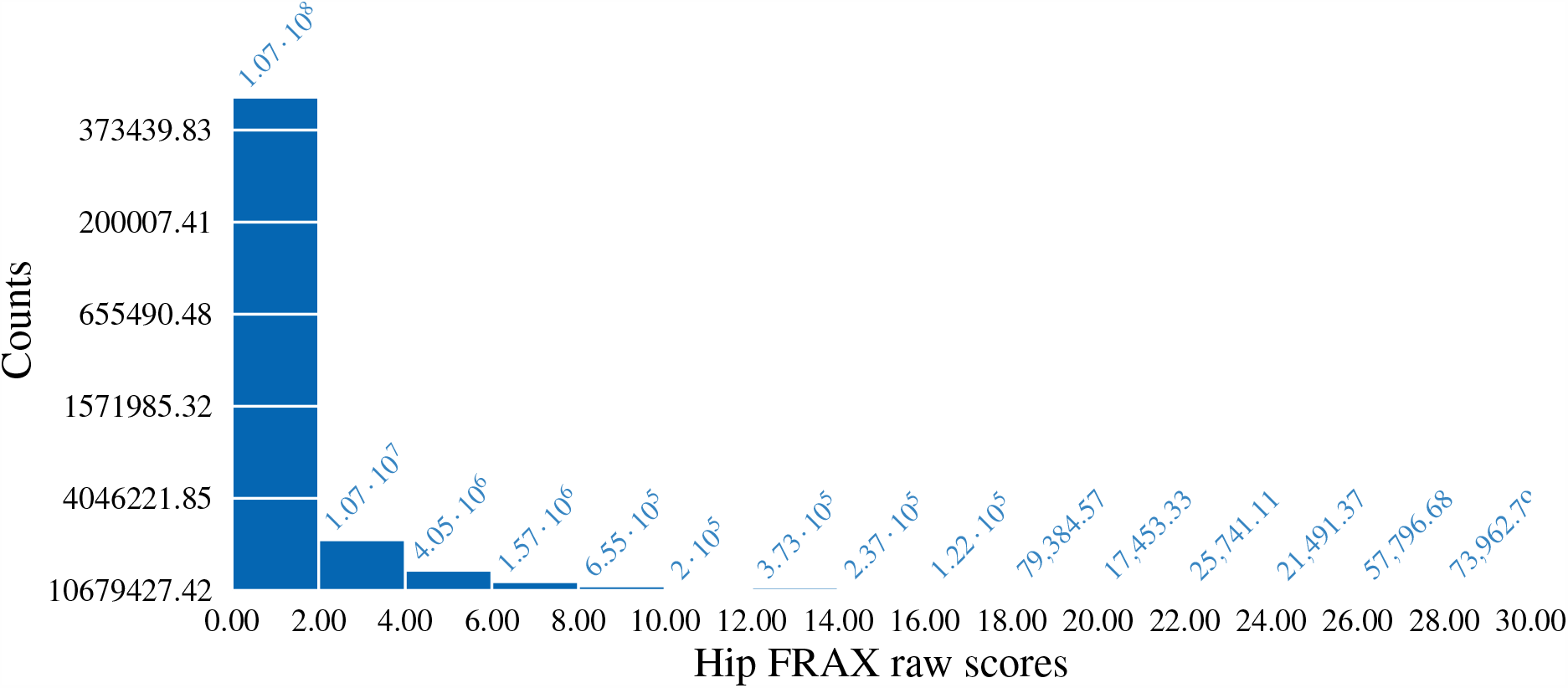
Histogram of hip FRAX raw scores

**Figure 6.**
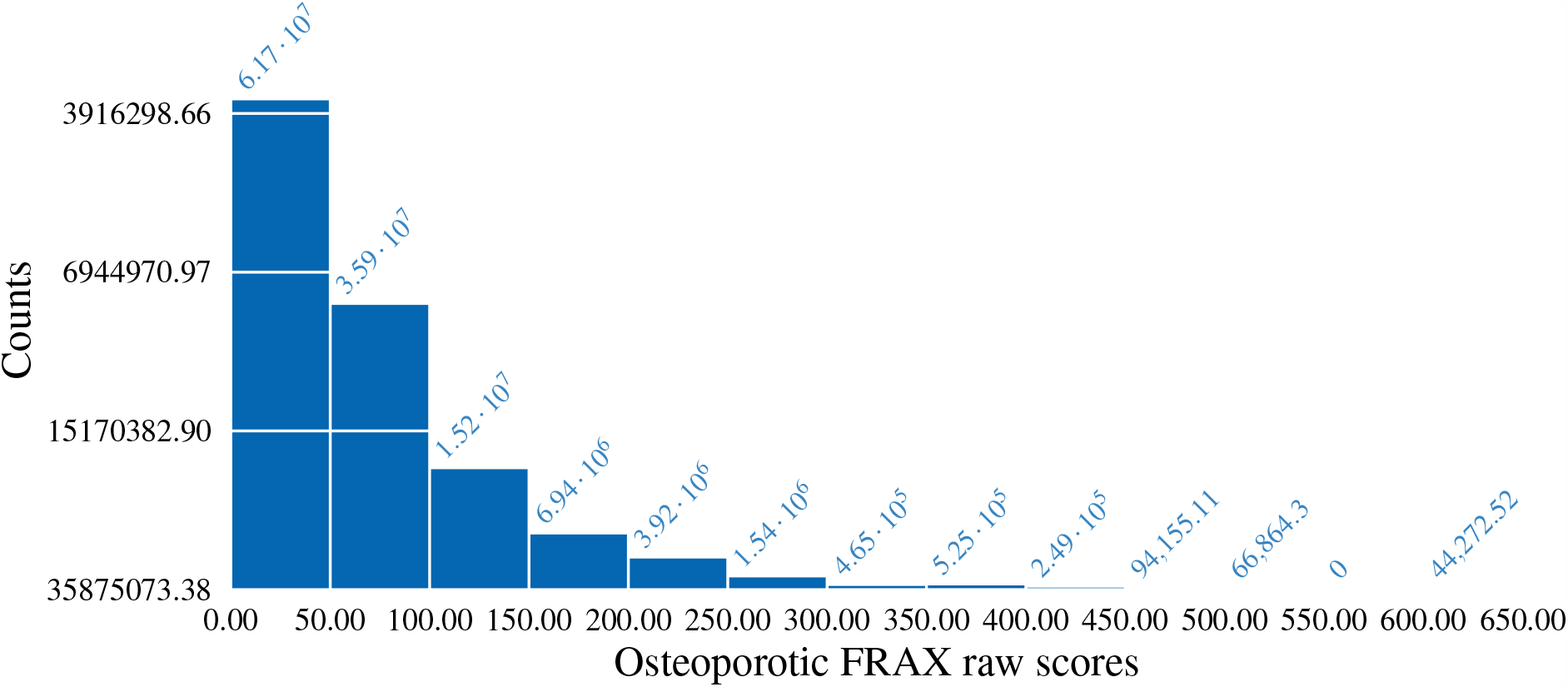
Histogram of osteoporotic FRAX raw scores

For hypothesis 3, clinical AL is the DV. This variable is available for 2011–14, and is calculated from raw data on probing depth and gingival recession by direct subtraction, with recession coded as negative when needed to get the math straight (Tomar and Asma, 2000). PD is diagnosed based on threshold criteria for both AL and pocket depth, but just AL may be a better measure of PD risk as opposed to prevalence. In NHANES 2009–10 periodontitis severity held steady with age, but AL severity rose. As risk is known to rise with age, AL with no thresholding seems the better measure for non-diagnostic purposes (Eke et al., 2012). Total AL is problematic because not all examined subjects have values for all sites. Hence, we must use the mean. Next, the variable is rounded to the nearest 0.1 to render it discrete. It now displays a Poisson shape (*Fig. 7*). There is no zero inflation (zero pockets for the whole mouth are rare), mean does not equal variance, and so the right regression call is svy vglm with negbinomial.

**Figure 7.**
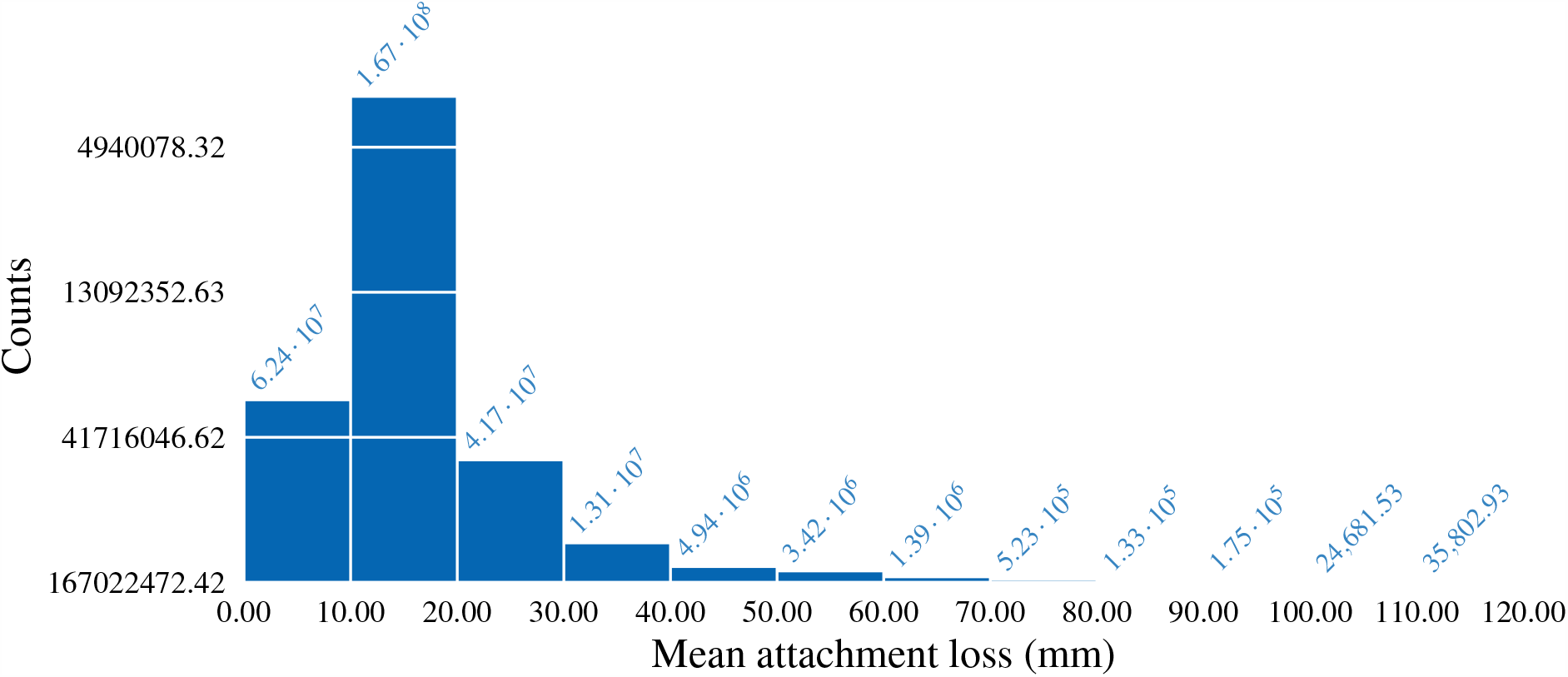
Histogram of mean attachment loss measures

### IVs

The IVs are all available for the years 2011–14. However, which years are used for a specific analysis depends on the DV. Clinical AL, associated with periodontitis, is the only DV spanning the whole range; the rest cover 2013–14.

#### 25(OH)D levels, LBXVIDMS

NHANES 2011–12 and 2013–14 measure just 25(OH)D serum levels. Further surveys dropped this measure, limiting all analyses at least to these years. Total 25(OH)D consists of VDBP bound 25(OH)D and free 25(OH)D. The free 25(OH)D gets converted eventually to the bioactive 1,25(OH)D, largely in the kidneys; the bound part is likely inert. Nevertheless, we are justified in using total 25(OH)D as our marker; studies such as the one by Zhang et al. (2019) have found the two to be correlated. That particular study, using WHI-OS data, found a correlation of *r*^2^ = 0.45 between total 25(OH)D and free 25(OH)D and *r*^2^ = 0.99 between free and bioavailable 25(OH)D. There is no biological significance to quantiles; hence this study uses the raw 25(OH)D level with no thresholding or corrections. No analysis in this study uses 25(OH)D as the DV; so, converting it to binary form is not required. The specific unit is nmol/L but that matters little as the other main unit, ng/mL, is just a scaling of this value.

Vitamin D and metabolite levels fluctuate by the seasons, particularly in the northern latitudes, likely because of variation in sunlight and sunlight exposure. In their MESA analysis, Robinson-Cohen et al. (2013) used a sine-wave to model monthly fluctuations in sample-average 25(OH)D and used that model to calculate an annual mean 25(OH)D value from the single value and month-of-blood-extraction present in the original data. They found the same results as using just the raw value. Our analyses include the 6-month stretch when the examination was conducted, RIDEXMON, the only information present in NHANES, as a covariate. RIDEXMON is one for winter and two for summer. Early version of NHANES did include the month of collection, as the variable name indicates.

#### Other IVs

We use the variables for age, RIDAGEYR in file DEMO, and sex, RIAGENDR, as is, except that the values in RIAGENDR are changed from 1/2 to 0/1 for convenience, keeping the more numerous group as code 0. The race/ethnicity variable, RIDRETH1, is processed to pick up just non-Hispanic whites (recoded as 0) and non-Hispanic AAs (recoded as 1). Education level, LDL, HDL, fasting plasma glucose levels, and BMI are picked up as is from variable DMDECU2 in file DEMO, LBDHDD in HDL, LBDLDL in TRIGLY, LBXGLU in GLU, and BMXBMI in BMX respectively. Both diastolic and systolic BPs are coded as the mean of the four available readings in BPXDI 1,2,34/BPXSI 1,2,3,4 in files BPX. Cigarette smoking is coded as the number of cigarettes smoked the previous month, available as the product of SMD641 (number of days smoked) and SMD650 (average number smoked a day), both from file SMQ. Non-smokers are identified via variable SMQ040, with number smoked set to 0 for them.

### Weighting, point estimates, standard errors

NHANES has a stratified, clustered, 4-stage design: 1. primary sampling unit (PSU) sampling counties, 2. census block segments, 3. dwelling units, and 4. individuals. Data collection is every year but release every two years. The sample is meant to represent both the noninstitutionalized, resident, civilian population of the 50 states of the US plus Washington, D.C., and its demographic breakdowns, for our purposes, the non-Hispanic white and AA populations and sex and age stratifications. NHANES used the Census Bureau’s 2011 and 2013 one-year American Community Survey (ACS) demographic numbers as the reference (Johnson et al., 2014). To ensure that representativeness holds in the analysis, each sample must be weighted by a number provided in the data. The variable that holds the weights varies. For interview data, the variable is WTINT2YR; for lab and physical exam data, it is WTMEC2YR and for fasting data, in our case plasma glucose and LDL levels, it is WTSAF2YR. In any analysis, the weights to be used are the most restrictive of all the variables in the analysis, with the most restrictive corresponding to the least frequent. Thus, WTSAF2YR is the most restrictive and WTMEC2YR next. We do not use WTINT2YR as no analysis has just interview data (Chen et al., 2018).

Point estimates, such as mean or variance of data or regression coefficients, must be adjusted to reflect the survey design. This is achieved by using the variables SDMVPSU and SDMVSTRA in the DEMO file along with the weights (Chen et al., 2018). SDMVPSU may not be unique across strata. Sampling is assumed to be with replacement. The R variable survey.lonely.psu is set to “adjust,” a conservative correction that centers a stratum with just one PSU at the grand mean instead of the stratum mean. If this were a certainty PSU the variance should be 0; the correction overestimates the variance here and is hence conservative. The variance estimator for the regression coefficients is the default provided by the R svyVGAM package, which is a Horvitz-Thompson-type sandwich estimator. This ensures the standard error of the coefficient is largely independent of the constant-variance assumption for error at different values, and that the stratified design is accounted for.

## RESULTS

For hypothesis 1 with AAC24 as the DV and race/ethnicity as the IV, the regression coefficient is significant at *p* < .001, with AAs having lower levels. Levels of 25(OH)D also show significance in another standalone regression, at *p* < 0.001, with more 25(OH)D leading to more AAC. But when the two IVs are combined with an interaction, only 25(OH)D is significant, at *p* < .01. *Table 1* shows the results in point-estimate/std-error form from a combined model, with most variables significant at at least *p* < .01, and sex significant at *p* < .05 with males having more AAC. In this analysis, the race/ethnicity coefficient is not significant, nor is the income to FPL ratio or systolic BP. When stratified by race, the 25(OH)D coefficient is no longer significant in AAs (*Table 2*) but is directly related at *p* < .01 in white Americans, that is, with more vitamin D leading to more AAC (*Table 3*).

**Table 2.**
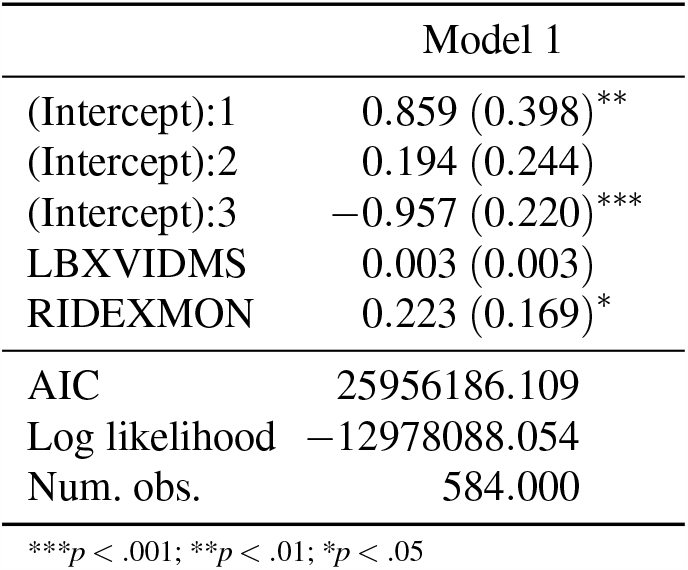
AAC vs vit D: nHBlacks only

|  | Model 1 |
| --- | --- |
| (Intercept):1 | 0.859 (0.398)** |
| (Intercept):2 | 0.194 (0.244) |
| (Intercept):3 | −0.957 (0.220)*** |
| LBXVIDMS | 0.003 (0.003) |
| RIDEXMON | 0.223 (0.169)* |
| AIC | 25956186.109 |
| Log likelihood | −12978088.054 |
| Num. obs. | 584.000 |

\*\*\* $p < .001$ ; \*\* $p < .01$ ; \* $p < .05$

**Table 3.** AAC vs vit D: nHwhites only

|  | Model 1 |
| --- | --- |
| (Intercept):1 | 1.257 (0.190)*** |
| (Intercept):2 | 0.152 (0.137) |
| (Intercept):3 | −0.529 (0.132)*** |
| LBXVIDMS | 0.003 (0.001)** |
| RIDEXMON | −0.025 (0.097) |
| AIC | 247308099.540 |
| Log likelihood | −123654044.770 |
| Num. obs. | 1350.000 |

\*\*\* $p < .001$ ; \*\* $p < .01$ ; \* $p < .05$

For hypothesis 2 with SBMD as the DV, the vitamin D coefficient is significant standalone at *p* < 0.01, with more vit. D decreasing BMD. When vit. D and race/ethnicity are combined in a model, only race/ethnicity stays significant at *p* < 0.001, with AAs having more SBMD. In the combined model of *Table 4*, no significance is seen for most variables. When the data are stratified by race/ethnicity, the vit. D coefficient is no longer significant in either case. With FBMD as the DV, the vit. D coefficient is significant at *p* < 0.001 in the standalone model with more vit. D leading to lower FBMD. When combined with race/ethnicity, it still stays significant at the same level, but race/ethnicity shows no significance. In the complete combined model of *Table 5*, race still has the same effect. When stratified by race/ethnicity, vit. D is significant in AAs at *p* < .05 (*Table 6*) and in white Americans at *p* < .001 (*Table 7*).

**Table 4.** SBMD vs race, vit D, smoking, income, edu., BMI, LDL, glucose, sys. BP, age, sex

|  | Model 1 |
| --- | --- |
| (Intercept) | 0.952 (0.126)** |
| RIDRETH1 | 0.057 (0.027) |
| blood_pressure_systole | -0.000 (0.001) |
| LBDLDL | -0.000 (0.000)* |
| DMDEDUC2 | 0.002 (0.010) |
| LBXVIDMS | 0.000 (0.000) |
| RIDEXMON | 0.016 (0.017) |
| BMXBMI | 0.009 (0.002)** |
| cig_smoking | -0.000 (0.000) |
| LBXGLU | -0.001 (0.000) |
| INDFMPIR | 0.002 (0.009) |
| RIDAGEYR | -0.003 (0.001) |
| RIAGENDR | 0.045 (0.034) |
| Deviance | 5.753 |
| Dispersion | 0.023 |
| Num. obs. | 229 |

\*\*\* $p < .001$ ; \*\* $p < .01$ ; \* $p < .05$

**Table 5.**
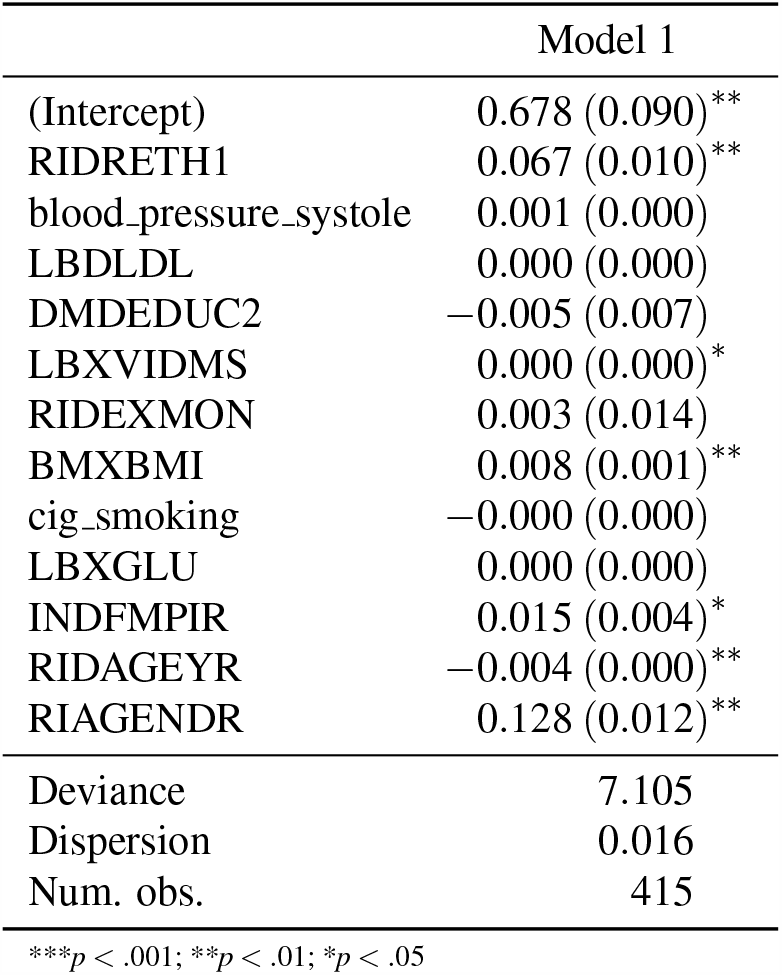
FBMD vs race, vit D, smoking, income, edu., BMI, LDL, glucose, sys. BP, age, sex

**Table 6.** FBMD vs vit D: nHBlacks only

|  | Model 1 |
| --- | --- |
| (Intercept) | 1.070 (0.030)*** |
| LBXVIDMS | -0.001 (0.000)* |
| RIDEXMON | -0.020 (0.018) |

| Model 1 |  |
| --- | --- |
| Deviance | 15.667 |
| Dispersion | 0.027 |
| Num. obs. | 590 |

\*\*\* $p < .001$ ; \*\* $p < .01$ ; \* $p < .05$

**Table 7.**
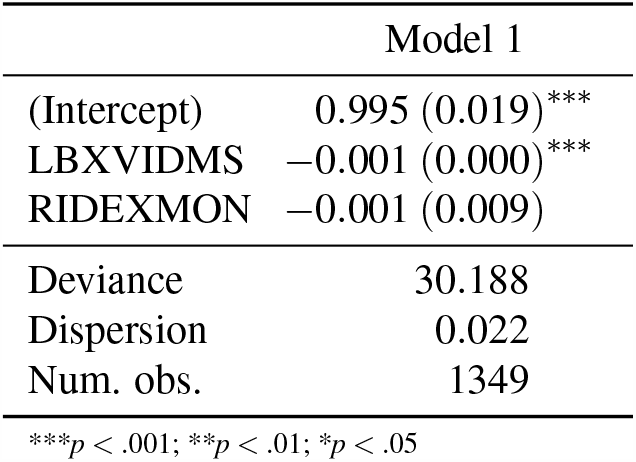
FBMD vs vit D: nHwhites only

With hip FRAX as the DV, vitamin D shows significance at *p* < .01 in a standalone model with higher levels of 25(OH) raising the FRAX score; combined with race/ethnicity and an interaction, the only significant coefficient is that of vit. D, at *p* < .05. A combined model shows age as significant at *p* < .01, with the risk rising with age. In stratified models, vitamin D stays significant, at *p* < .05, for white Americans (*Table 9*) but not for AAs (*Table 8*). With osteoporotic FRAX as the DV, a standalone model shows both vitamin D as significant, at *p* < .001 and the season as well, at *p* < .01. Higher vitamin D and winter are protective. *Table 10* shows the results of a combined model. Almost all variables are significant, except for season. In stratified models with all control variables added, vit. D is significant for just white Americans at *p* < .001 (*Table 11* and *Table 12*).

**Table 8.** Hip fracture risk vs vit D: nHBlacks only

| Model 1 |  |
| --- | --- |
| (Intercept) | −0.006 (0.118) |
| LBXVIDMS | 0.003 (0.001) |
| RIDEXMON | 0.119 (0.062) |
| Deviance | 554.332 |
| Dispersion | 0.935 |
| Num. obs. | 594 |

\*\*\* $p < .001$ ; \*\* $p < .01$ ; \* $p < .05$

**Table 9.** Hip fracture risk vs vit D: nHwhites only

| Model 1 |  |
| --- | --- |
| (Intercept) | 1.008 (0.321)** |
| LBXVIDMS | 0.005 (0.002)* |
| RIDEXMON | 0.065 (0.171) |
| Deviance | 9352.711 |
| Dispersion | 6.949 |
| Num. obs. | 1347 |

\*\*\* $p < .001$ ; \*\* $p < .01$ ; \* $p < .05$

**Table 10.** Osteo. frac. risk vs vit D, race, syst. BP, LDL, edu., BMI, smoking, glucose, income ratio, age, sex

| Model 1 |  |
| --- | --- |
| (Intercept):1 | 3.098 (0.039)*** |
| (Intercept):2 | 1.458 (0.044)*** |

|  | Model 1 |
| --- | --- |
| RIDRETH1 | −1.177 (0.022)*** |
| blood_pressure_systole | −0.001 (0.000)*** |
| LBDLDL | −0.000 (0.000)* |
| DMDEDUC2 | 0.068 (0.018)*** |
| LBXVIDMS | −0.002 (0.000)*** |
| RIDEXMON | 0.008 (0.011) |
| BMXBMI | −0.009 (0.003)*** |
| cig_smoking | 0.000 (0.000)*** |
| LBXGLU | −0.001 (0.000)*** |
| INDFMPIR | −0.075 (0.008)*** |
| RIDAGEYR | 0.037 (0.000)*** |
| RIAGENDR | −0.474 (0.009)*** |
| AIC | 447609036.742 |
| Log likelihood | −223804504.371 |
| Num. obs. | 446.000 |

\*\*\* $p < .001$ ; \*\* $p < .01$ ; \* $p < .05$

**Table 11.**
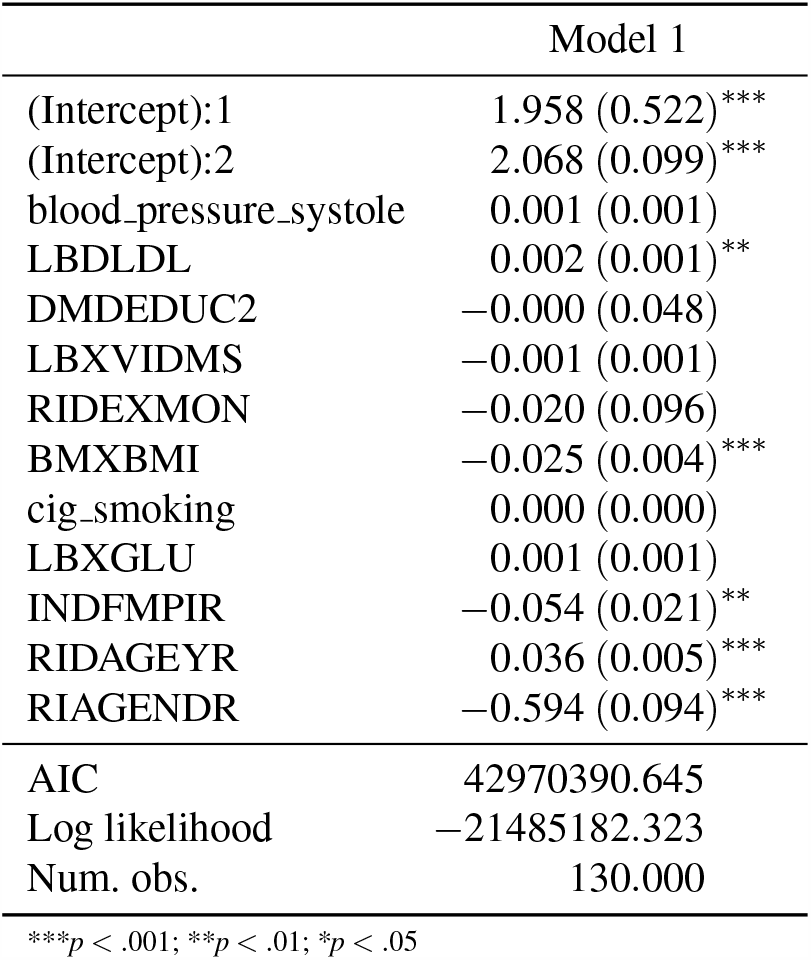
Osteo. frac. risk vs vit D, syst. BP, LDL, edu., BMI, smoking, glucose, income ratio, age, sex: nHBlack

|  | Model 1 |
| --- | --- |
| (Intercept):1 | 1.958 (0.522)*** |
| (Intercept):2 | 2.068 (0.099)*** |
| blood_pressure_systole | 0.001 (0.001) |
| LBDLDL | 0.002 (0.001)** |
| DMDEDUC2 | −0.000 (0.048) |
| LBXVIDMS | −0.001 (0.001) |
| RIDEXMON | −0.020 (0.096) |
| BMXBMI | −0.025 (0.004)*** |
| cig_smoking | 0.000 (0.000) |
| LBXGLU | 0.001 (0.001) |
| INDFMPIR | −0.054 (0.021)** |
| RIDAGEYR | 0.036 (0.005)*** |
| RIAGENDR | −0.594 (0.094)*** |
| AIC | 42970390.645 |
| Log likelihood | −21485182.323 |
| Num. obs. | 130.000 |

\*\*\* $p < .001$ ; \*\* $p < .01$ ; \* $p < .05$

**Table 12.** Osteo. frac. risk vs vit D, syst. BP, LDL, edu., BMI, smoking, glucose, income ratio, age, sex: nHwhite

|  | Model 1 |
| --- | --- |
| (Intercept):1 | 3.122 (0.119)*** |
| (Intercept):2 | 1.416 (0.028)*** |
| blood_pressure_systole | −0.001 (0.001)* |
| LBDLDL | −0.000 (0.001) |
| DMDEDUC2 | 0.073 (0.033)** |
| LBXVIDMS | −0.002 (0.000)*** |
| RIDEXMON | 0.004 (0.012) |
| BMXBMI | −0.007 (0.002)*** |
| cig_smoking | 0.000 (0.000)*** |
| LBXGLU | −0.002 (0.000)*** |
| INDFMPIR | −0.075 (0.013)*** |
| RIDAGEYR | 0.037 (0.002)*** |
| RIAGENDR | −0.462 (0.020)*** |
| AIC | 403460117.597 |
| Log likelihood | −201730045.798 |
| Num. obs. | 316.000 |

\*\*\* $p < .001$ ; \*\* $p < .01$ ; \* $p < .05$

For hypothesis 3, the only DV is AL. *Table 13* shows the results of a combined model with all variables except LDL, systolic BP, and season significant, largely at *P* < .001 with GLU at *p* < .01. More vitamin D reduces AL; AAs have more AL and so do males, the older, the poorer, the smoker, the less educated, and counter intuitively those with lower BMI. As *Table 14* and *Table 15* indicate, stratification by race/ethnicity keeps vit. D significant, at *p* < .01 for AAs and at *p* < .05 for white Americans, after the addition of all our control variables.

**Table 13.** AL vs vit D, race, systolic BP, LDL, edu., BMI, smoking, glucose, income ratio, age, sex

|  | Model 1 |
| --- | --- |
| (Intercept):1 | 2.619 (0.085)*** |
| (Intercept):2 | 1.403 (0.035)*** |
| RIDRETH1 | 0.143 (0.046)*** |
| blood_pressure_systole | 0.002 (0.002) |
| LBDLDL | 0.001 (0.001) |
| DMEDEDUC2 | −0.155 (0.021)*** |
| LBXVIDMS | −0.004 (0.000)*** |
| RIDEXMON | 0.001 (0.050) |
| BMXBMI | −0.010 (0.001)*** |
| cig_smoking | 0.000 (0.000)*** |
| LBXGLU | 0.001 (0.001)** |
| INDFMPIR | −0.072 (0.010)*** |
| RIDAGEYR | 0.016 (0.002)*** |
| RIAGENDR | 0.215 (0.028)*** |
| AIC | 334280231.303 |
| Log likelihood | −167140101.652 |
| Num. obs. | 869.000 |

\*\*\* $p < .001$ ; \*\* $p < .01$ ; \* $p < .05$

**Table 14.** AL vs vit D, systolic BP, LDL, edu., BMI, smoking, glucose, income ratio, age, sex: nHBlack only

|  | Model 1 |
| --- | --- |
| (Intercept):1 | 2.398 (0.199)*** |
| (Intercept):2 | 1.617 (0.101)*** |
| blood_pressure_systole | 0.003 (0.002)* |
| LBDLDL | −0.000 (0.001) |
| DMEDEDUC2 | −0.118 (0.023)*** |
| LBXVIDMS | −0.004 (0.002)** |
| RIDEXMON | 0.042 (0.062) |
| BMXBMI | −0.008 (0.006)* |
| cig_smoking | −0.000 (0.000)* |
| LBXGLU | −0.001 (0.001) |
| INDFMPIR | −0.016 (0.028) |
| RIDAGEYR | 0.019 (0.002)*** |
| RIAGENDR | 0.230 (0.077)*** |
| AIC | 48001327.311 |
| Log likelihood | −24000650.656 |
| Num. obs. | 275.000 |

\*\*\* $p < .001$ ; \*\* $p < .01$ ; \* $p < .05$

**Table 15.** AL vs vit D, systolic BP, LDL, edu., BMI, smoking, glucose, income ratio, age, sex: nHwhite only

|  | Model 1 |
| --- | --- |
| (Intercept):1 | 2.325 (0.363)*** |
| (Intercept):2 | 1.355 (0.130)*** |
| blood_pressure_systole | 0.002 (0.004) |
| LBDLDL | 0.000 (0.001) |
| DMDEDUC2 | −0.154 (0.040)*** |
| LBXVIDMS | −0.003 (0.002)* |
| RIDEXMON | −0.013 (0.059) |
| BMXBMI | −0.008 (0.003)** |
| cig_smoking | 0.001 (0.000)*** |
| LBXGLU | 0.002 (0.001)* |
| INDFMPIR | −0.067 (0.032)** |
| RIDAGEYR | 0.018 (0.002)*** |
| RIAGENDR | 0.213 (0.047)*** |
| AIC | 281764025.080 |
| Log likelihood | −140881999.540 |
| Num. obs. | 594.000 |

\*\*\* $p < .001$ ; \*\* $p < .01$ ; \* $p < .05$

## DISCUSSION

### Hypothesis 1

Hypothesis 1 is confirmed by the data, though with several niceties. AAC24 does depend on vit. D, with more vit. D raising AAC scores in white Americans, with AAs showing no effect. However, the lack of an effect in AAs is due to higher standard errors for the coefficients, presumably from the smaller sample size. Absence of proof in this case does not point to proof of likely absence. In the overall populace, the race/ethnicity effect vanishes when vit. D is added to the regression. This means lower vit. D is protective when it comes to AAC; the race/ethnicity difference is due to lower vit. D levels in AAs. Incidentally, the inverse effects for BMI and LDL on AAC24 in the overall data are hard to explain. The data also make it appear that AAC24 scores are lower in summer; this is probably due to the geographic variation in where NHANES samples data in summer and winter. As the full model with all our IVs thrown in shows no race/ethnicity influence on AAC24, we must conclude that lower vit. D, in and of itself, is protective for AAC. Further studies are required to see whether this result applies to CAC as well. A mechanistic interpretation is that higher 25(OH)D facilitates higher calcium absorption in the intestines and deposition of that calcium elsewhere. However, this finding contrasts with others that found low 25(OH)D to produce more CAC in white Americans.

### Hypothesis 2

The second hypothesis was only partly validated. In sum, higher levels of 25(OH)D reduced FBMD in both groups, increased hip FRAX in white Americans only, and decreased osteoporotic FRAX in white Americans. The effects of 25(OH)D on osteoporotic FRAX was similar in white Americans and AAs after stratification with no control variables: in both cases higher vit. D led to higher risk of osteoporotic fractures, possibly through reducing BMD. Addition of all the control variables led to an inversion of the relationship in white Americans, with higher vit. D leading to lower osteoporosis risk. In AAs, statistical significance was lost, presumably due to the smaller sample size. The results for osteoporotic FRAX held in white Americans after adding controls for systolic BP, education, BMI, smoking, fasting plasma glucose levels, income to FPL ratio, age, and sex, along with LDL and season, both nonsignificant. The increased hip FRAX with vit. D for white Americans is likely an artifact; adding all the control variables made the effect vanish (result not shown). To recapitulate, vitamin D increase reduces BMD in both groups but reduces FRAX just in white Americans. In AAs, with no controls and hence a larger sample, 25(OH)D rise increases FRAX; this effect vanishes when controls are added. This may be because the controls do explain the 25(OH)D link or because the sample size gets reduced.

### Hypothesis 3

Hypothesis 3 was borne out unequivocally. Increased vit. D levels reduce periodontal risk in white Americans, AAs, and the total sample. The results are robust after controlling for systolic BP, LDL, education, season, BMI, cigarette smoking, plasma glucose level, income to FPL ratio, age, and sex. All those variables except for BP, LDL, and season show relationships with AL in white Americans; in AAs, the above plus fasting plasma glucose and income/FPL show no relationship, all else does. BMI is a surprise, with higher BMI leading to lower AL scores. Males seemingly are more at risk than females, *p* < .001 in both groups; this coefficient is in fact the largest in both stratified models and the combined model (as units differ, coefficients are not directly comparable, but as the unit distance is the largest, 0 to 1, for sex, this high coefficient is all the more remarkable). In the combined model, AAs are significantly at more risk after all the additional variables are controlled for. This may indicate residual risk from ancestral genetics but more so from unmodeled environmental elements such as stress and unequal receipt of treatment even after controlling for socio-economic status.

### Differential impact of vit. D

For both aortic calcification, at least in the abdominal aorta, and for bone health, measured by FRAX scores and BMDs, the influence of vit. D is stratified by race/ethnicity. The alternative, that the better numbers in AAs for both elements is from protective elements overriding the vit. D deficiency effect finds no support in this analysis. The various physiological, biochemical, and environmental factors included in the controls do not exhaust all possibilities but do cover enough ground to conclude that the vit. D effect is a direct one. Many unmodeled environmental elements such as stress are not likely to be protective in AAs.

Periodontitis risk shows no such race/ethnicity effect; the risk is inversely proportional to vit. D levels across the board, once again with all the controls added. This does argue for the risk in AAs being directly from vit. D deficiency. One notes that the unmodeled environmental elements may weaken that conclusion. However, stress and such psychosocial elements are not easy to measure quantitatively and hence are likely to be out of the purview of survey analysis.

We thus see two diametrically opposite conclusions for CVD/bone health and oral health. For the first set, differing vit. D thresholds for different groups do look warranted. As race and ethnicity are both social constructs, how exactly to construct such differing thresholds is a challenge. Most likely, the assessment has to be individualized, based on ancestral genetic markers, context of the living environment, dietary elements, and the like. A single threshold for the entire populace has negative relevance for the CVD risk and bone health elements of AAs, as their risks rise with vit. D levels. The periodontitis data argue against this conclusion. However, PD is less of a burden on quality of life and longevity than CVD or fractures. PD is also more avoidable with improved and admittedly more inconvenient dental hygiene. Vitamin D level recommendations must hence probably focus more on CVD and fracture risks as the relatively greater risks.

This analysis, like most others in the literature, cannot extract causality from the data. NHANES provides cross-sectional data and so only correlations between 25(OH)D and other variables can be estimated. In some cases, such as gum inflammation, there is some evidence that the disease increases local production of vit. D. In CVD and bone fracture risks, there is a possibility that some other element contributes to both vit. D levels and the risk factor. Physiological markers such as blood pressure and BMI; biochemical markers such as fasting plasma glucose level and LDL level; climatic elements such as the season; the demographic factors of age and sex; and socioeconomic variables such as income to FPL ratio and educational level, are all included in this analysis to account for the influence of such hidden variables. The central conclusion of inverse relation across races/ethnicities between 25(OH)D and CVD and bone health risk holds even with all those variables controlled for, and after stratification of data by the groups in question.

Vitamin D levels can be mapped via multiple biomarkers. Free 25(OH)D is believed to be what gets converted to the bioactive 1,25(OH)D form. NHANES, in common with most other surveys, used total 25(OH)D as a measure of vitamin D levels. This marker has been standardized, initially by the US National Institute of Standards, and then by Ghent University, Belgium (Brown et al., 2018). No such reference exists for other markers. The NHANES data used in this study conforms to the reference standard. Free and total BMD are highly correlated Zhang et al. (2019), and hence perhaps this issue is not as relevant as it appears.

The vit. D paradox in AAs has been known for some time. Brown et al. (2018), a report on an expert panel meeting, notes that overall mortality was strongly and inversely linked to 25(OH)D for white Americans but not so for AAs in the 1988 to 2010 NHANES data. The report also mentions that the MESA trial showed lower fracture rates overall for AAs, and a direct relationship between CVD risk and 25(OH)D in AAs as opposed to an inverse relationship in white Americans and some other groups. This study confirms that finding in the 2011–14 NHANES data. Low vit. D levels can lead to osteomalacia in adults, but the level at which the protective effect levels off is 12 ng/mL, well below the recommendation of 30 ng/mL. The expert panel report explicitly notes that the “higher levels of vitamin D [present] hazards for Black Americans, resulting in increasing falls and fractures” (Brown et al., 2018, p. 11). This study confirms the fracture risk increase and general bone health issues, such as decreasing BMD, with increasing 25(OH)D levels in AAs.

For clinical recommendations, information on other effects of vitamin D levels must also be considered, specifically its effects on cancer incidence and growth, reproductive health, metabolic syndrome, COPD, acute respiratory distress, and the general immune system. Nevertheless, the specific harm implied by the statistical relation between higher serum 25(OH)D and worse bone and cardiovascular health in AAs merits a closer look. NHANES no longer collects vit. D levels. Directly assessing the relationship between vit. D levels and markers of the health and illness conditions mentioned earlier requires a nationally representative sample with the scale and scope of NHANES.

## CONCLUSIONS

This analysis finds that higher levels of 25(OH)D may be harmful to AAs as far as abdominal aortic calcification, bone mineral density, and osteoporotic fracture risks go but protective with regards to oral, specifically periodontal, health. In the case of AAC, higher vit. D levels are harmful for white Americans, with statistical significance lacking for a similar relationship in AAs; for bone health the relationship is inverted for the two groups with higher 25(OH)D benefiting white Americans and harming AAs, specifically with regards to BMD and for models with standalone FRAX scores. For periodontal risk, attachment loss rises with lower levels of 25(OH)D for both groups. The control variables used in these analyses are LDL levels, fasting plasma glucose levels, systolic BP, BMI, age, sex, income to federal poverty level ratio, educational level, and season of blood extraction. These results must be complemented with collection and analysis of data on other health effects of vit. D to figure out appropriate vit. D recommendations for AAs in particular, and likely other groups as well, based on a holistic and individualized assessment.

## Data Availability

Publicly available data from the CDC (the NHANES study).

https://www.cdc.gov/nchs/nhanes/index.htm

---

**Supplemental Table 1.** AAC vs race

| Model 1 |  |
| --- | --- |
| (Intercept):1 | 1.490 (0.059)*** |
| (Intercept):2 | 0.152 (0.124) |
| (Intercept):3 | −0.580 (0.122)*** |
| RIDRETH1 | −0.252 (0.065)*** |
| AIC | 279596429.431 |
| Log likelihood | −139798210.716 |
| Num. obs. | 1995.000 |

\*\*\* $p < .001$ ; \*\* $p < .01$ ; \* $p < .05$

**Supplemental Table 2.**
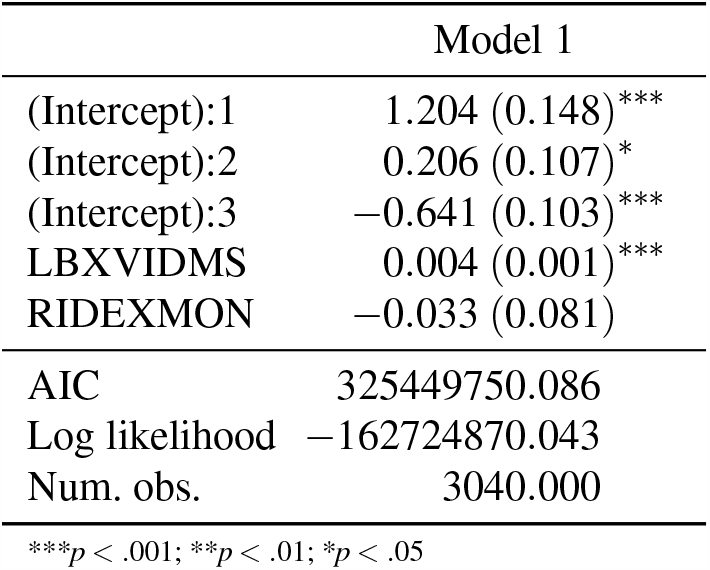
AAC vs vit D

**Supplemental Table 3.** AAC vs vit D, race, interaction

| Model 1 |  |
| --- | --- |
| (Intercept):1 | 1.237 (0.185)*** |
| (Intercept):2 | 0.139 (0.119) |
| (Intercept):3 | −0.567 (0.120)*** |
| LBXVIDMS | 0.003 (0.001)** |
| RIDEXMON | −0.006 (0.093) |
| RIDRETH1 | −0.238 (0.206) |
| LBXVIDMS:RIDRETH1 | 0.000 (0.003) |
| AIC | 273552474.277 |
| Log likelihood | −136776230.138 |
| Num. obs. | 1934.000 |

\*\*\* $p < .001$ ; \*\* $p < .01$ ; \* $p < .05$

**Supplemental Table 4.**
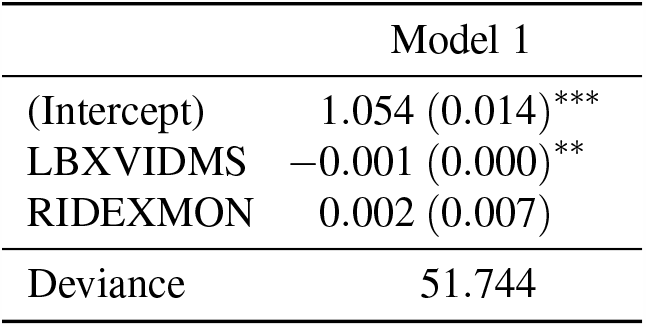
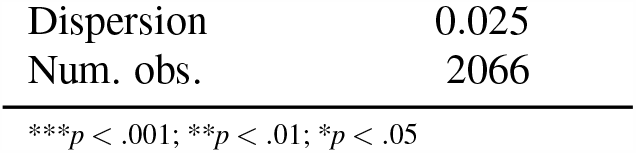
SBMD vs vit D

| Model 1 |  |
| --- | --- |
| (Intercept) | 1.054 (0.014)*** |
| LBXVIDMS | −0.001 (0.000)** |
| RIDEXMON | 0.002 (0.007) |
| Deviance | 51.744 |
| Dispersion | 0.025 |
| Num. obs. | 2066 |

\*\*\* $p < .001$ ; \*\* $p < .01$ ; \* $p < .05$

**Supplemental Table 5.** SBMD vs vit D, race

| Model 1 |  |
| --- | --- |
| (Intercept) | 1.037 (0.017)*** |
| LBXVIDMS | −0.000 (0.000) |
| RIDEXMON | 0.005 (0.008) |
| RIDRETH1 | 0.066 (0.009)*** |
| Deviance | 39.496 |
| Dispersion | 0.032 |
| Num. obs. | 1223 |

\*\*\* $p < .001$ ; \*\* $p < .01$ ; \* $p < .05$

**Supplemental Table 6.** SBMD vs vit D, race, interaction

| Model 1 |  |
| --- | --- |
| (Intercept) | 1.037 (0.018)*** |
| LBXVIDMS | −0.000 (0.000) |
| RIDEXMON | 0.005 (0.008) |
| RIDRETH1 | 0.067 (0.021)** |
| LBXVIDMS:RIDRETH1 | −0.000 (0.000) |
| Deviance | 39.496 |
| Dispersion | 0.032 |
| Num. obs. | 1223 |

\*\*\* $p < .001$ ; \*\* $p < .01$ ; \* $p < .05$

**Supplemental Table 7.**
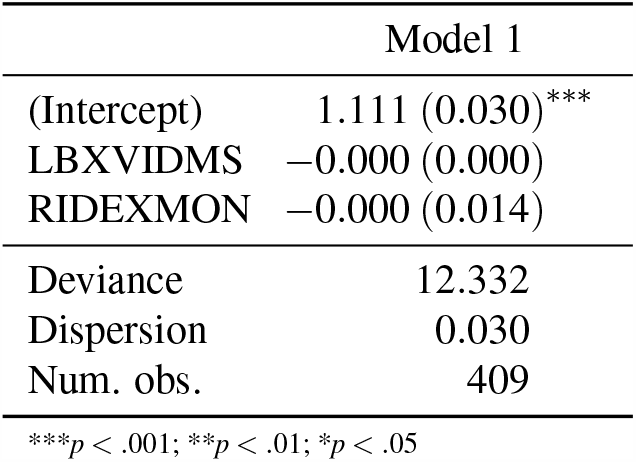
SBMD vs vit D: nHBlacks only

| Model 1 |  |
| --- | --- |
| (Intercept) | 1.111 (0.030)*** |
| LBXVIDMS | −0.000 (0.000) |
| RIDEXMON | −0.000 (0.014) |
| Deviance | 12.332 |
| Dispersion | 0.030 |
| Num. obs. | 409 |

\*\*\* $p < .001$ ; \*\* $p < .01$ ; \* $p < .05$

**Supplemental Table 8.** SBMD vs vit D: nHwhites only

| Model 1 |  |
| --- | --- |
| (Intercept) | 1.036 (0.019)*** |
| LBXVIDMS | −0.000 (0.000) |
| RIDEXMON | 0.006 (0.009) |
| Deviance | 20.646 |
| Dispersion | 0.025 |
| Num. obs. | 814 |

\*\*\* $p < .001$ ; \*\* $p < .01$ ; \* $p < .05$

**Supplemental Table 9.** FBMD vs vit D

| Model 1 |  |
| --- | --- |
| (Intercept) | 1.018 (0.014)*** |
| LBXVIDMS | −0.001 (0.000)*** |
| RIDEXMON | −0.005 (0.008) |
| Deviance | 69.516 |
| Dispersion | 0.023 |
| Num. obs. | 3027 |

\*\*\* $p < .001$ ; \*\* $p < .01$ ; \* $p < .05$

**Supplemental Table 10.** FBMD vs vit D, race

| Model 1 |  |
| --- | --- |
| (Intercept) | 0.997 (0.017)*** |
| LBXVIDMS | −0.001 (0.000)*** |
| RIDEXMON | −0.004 (0.008) |
| RIDRETH1 | 0.059 (0.008)*** |
| Deviance | 56.545 |
| Dispersion | 0.029 |
| Num. obs. | 1939 |

\*\*\* $p < .001$ ; \*\* $p < .01$ ; \* $p < .05$

**Supplemental Table 11.** FBMD vs vit D, race, interaction

| Model 1 |  |
| --- | --- |
| (Intercept) | 0.998 (0.017)*** |
| LBXVIDMS | −0.001 (0.000)*** |
| RIDEXMON | −0.004 (0.008) |
| RIDRETH1 | 0.050 (0.019)* |
| LBXVIDMS:RIDRETH1 | 0.000 (0.000) |
| Deviance | 56.540 |
| Dispersion | 0.029 |
| Num. obs. | 1939 |

\*\*\* $p < .001$ ; \*\* $p < .01$ ; \* $p < .05$

**Supplemental Table 12.**
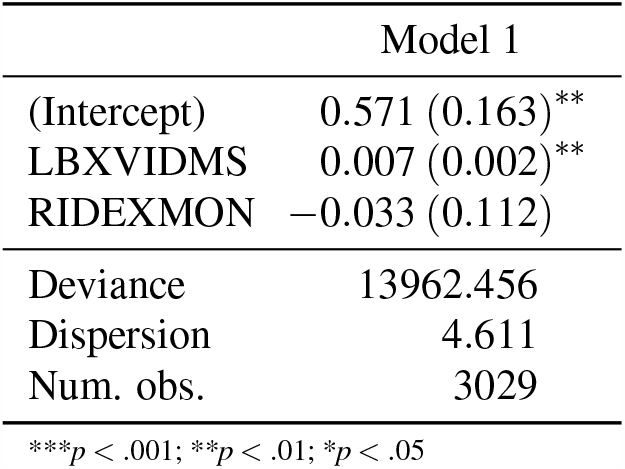
Hip fracture risk vs vit D

| Model 1 |  |
| --- | --- |
| (Intercept) | 0.571 (0.163)** |
| LBXVIDMS | 0.007 (0.002)** |
| RIDEXMON | -0.033 (0.112) |
| Deviance | 13962.456 |
| Dispersion | 4.611 |
| Num. obs. | 3029 |

\*\*\* $p < .001$ ; \*\* $p < .01$ ; \* $p < .05$

**Supplemental Table 13.**
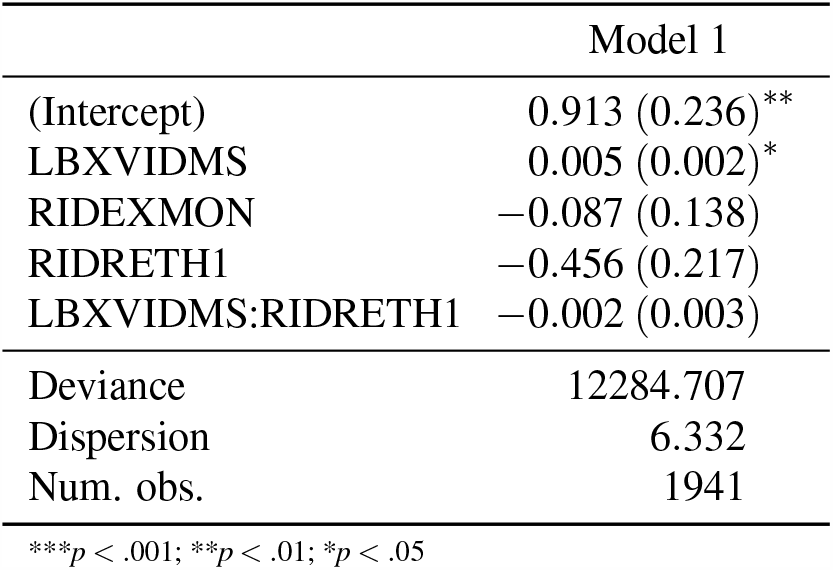
Hip fracture risk vs vit D, race, interaction

| Model 1 |  |
| --- | --- |
| (Intercept) | 0.913 (0.236)** |
| LBXVIDMS | 0.005 (0.002)* |
| RIDEXMON | -0.087 (0.138) |
| RIDRETH1 | -0.456 (0.217) |
| LBXVIDMS:RIDRETH1 | -0.002 (0.003) |
| Deviance | 12284.707 |
| Dispersion | 6.332 |
| Num. obs. | 1941 |

\*\*\* $p < .001$ ; \*\* $p < .01$ ; \* $p < .05$

**Supplemental Table 14.** Hip fracture risk vs vit D, race, systolic BP, LDL, edu., BMI, smoking, glucose, income ratio, age, sex

| Model 1 |  |
| --- | --- |
| (Intercept) | -0.934 (1.106) |
| RIDRETH1 | -0.648 (0.297) |
| blood_pressure_systole | 0.002 (0.004) |
| LBDLDL | -0.005 (0.005) |
| DMDEDUC2 | 0.180 (0.123) |
| LBXVIDMS | -0.004 (0.003) |
| RIDEXMON | -0.053 (0.257) |
| BMXBMI | -0.014 (0.016) |
| cig_smoking | 0.001 (0.001) |
| LBXGLU | -0.004 (0.002) |
| INDFMPIR | -0.071 (0.089) |
| RIDAGEYR | 0.055 (0.009)** |
| RIAGENDR | -0.060 (0.225) |
| Deviance | 2254.183 |
| Dispersion | 5.147 |
| Num. obs. | 408 |

\*\*\* $p < .001$ ; \*\* $p < .01$ ; \* $p < .05$

**Supplemental Table 15.** Osteo. fracture risk vs vit D

| Model 1 |  |
| --- | --- |
| (Intercept):1 | 3.716 (0.168)*** |
| (Intercept):2 | 0.541 (0.034)*** |
| LBXVIDMS | 0.006 (0.001)*** |
| RIDEXMON | 0.094 (0.051)* |
| AIC | 1286558524.981 |
| Log likelihood | −643279258.491 |
| Num. obs. | 3027.000 |

\*\*\* $p < .001$ ; \*\* $p < .01$ ; \* $p < .05$ **Supplemental Table 16.** Osteo. fracture risk vs vit D, race, interaction

**Supplemental Table 16.** Osteo. fracture risk vs vit D, race, interaction

| Model 1 |  |
| --- | --- |
| (Intercept):1 | 4.223 (0.134)*** |
| (Intercept):2 | 0.791 (0.026)*** |
| LBXVIDMS | 0.002 (0.001)** |
| RIDEXMON | 0.040 (0.049) |
| RIDRETH1 | −1.260 (0.125)*** |
| LBXVIDMS:RIDRETH1 | 0.002 (0.002) |
| AIC | 1046495101.316 |
| Log likelihood | −523247544.658 |
| Num. obs. | 1939.000 |

\*\*\* $p < .001$ ; \*\* $p < .01$ ; \* $p < .05$ **Supplemental Table 17.** Osteo. fracture risk vs vit D: nHBlacks only

**Supplemental Table 17.** Osteo. fracture risk vs vit D: nHBlacks only

| Model 1 |  |
| --- | --- |
| (Intercept):1 | 2.930 (0.163)*** |
| (Intercept):2 | 0.978 (0.117)*** |
| LBXVIDMS | 0.004 (0.001)*** |
| RIDEXMON | 0.065 (0.091) |
| AIC | 99306369.252 |
| Log likelihood | −49653180.626 |
| Num. obs. | 590.000 |

\*\*\* $p < .001$ ; \*\* $p < .01$ ; \* $p < .05$ **Supplemental Table 18.** Osteo. fracture risk vs vit D: nHwhites only

**Supplemental Table 18.**
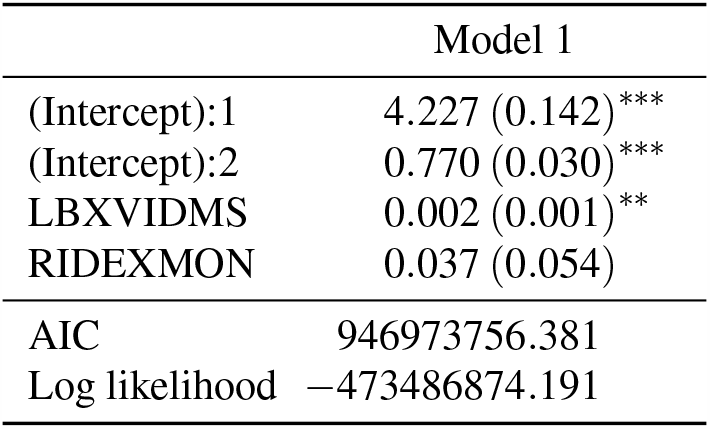
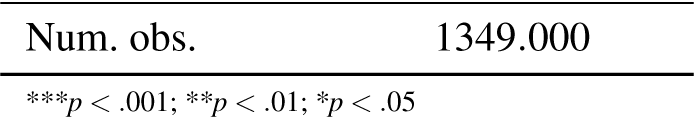
Osteo. fracture risk vs vit D: nHwhites only

**Supplemental Table 19.** AL vs vit D

|  | Model 1 |
| --- | --- |
| (Intercept):1 | 3.073 (0.071)*** |
| (Intercept):2 | 1.701 (0.054)*** |
| LBXVIDMS | −0.003 (0.000)*** |
| RIDEXMON | −0.041 (0.040) |
| AIC | 987249289.625 |
| Log likelihood | −493624640.812 |
| Num. obs. | 6675.000 |

\*\*\* $p < .001$ ; \*\* $p < .01$ ; \* $p < .05$

**Supplemental Table 20.**
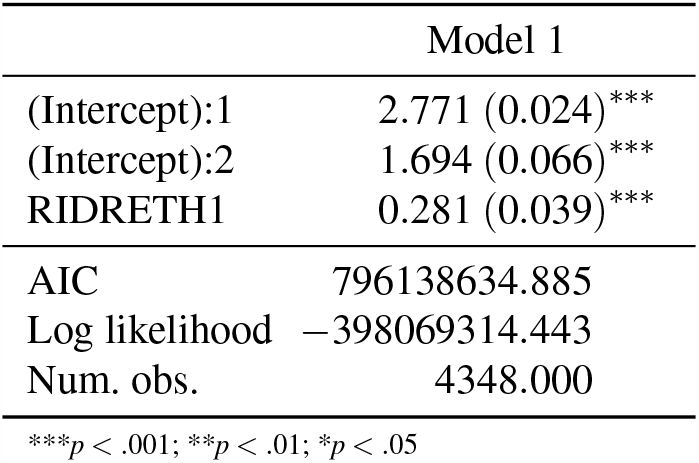
AL vs race

**Supplemental Table 21.** AL vs vit D, race

|  | Model 1 |
| --- | --- |
| (Intercept):1 | 2.980 (0.076)*** |
| (Intercept):2 | 1.715 (0.070)*** |
| RIDRETH1 | 0.231 (0.042)*** |
| LBXVIDMS | −0.002 (0.001)*** |
| RIDEXMON | −0.040 (0.043) |
| AIC | 771501488.418 |
| Log likelihood | −385750739.209 |
| Num. obs. | 4179.000 |

\*\*\* $p < .001$ ; \*\* $p < .01$ ; \* $p < .05$

**Supplemental Table 22.** AL vs vit D, race, interaction

|  | Model 1 |
| --- | --- |
| (Intercept):1 | 2.987 (0.078)*** |
| (Intercept):2 | 1.716 (0.070)*** |
| RIDRETH1 | 0.187 (0.062)*** |

| Model 1 |  |
| --- | --- |
| LBXVIDMS | −0.002 (0.001)*** |
| RIDEXMON | −0.039 (0.043) |
| RIDRETH1:LBXVIDMS | 0.001 (0.001) |
| AIC | 771473908.664 |
| Log likelihood | −385736948.332 |
| Num. obs. | 4179.000 |

\*\*\* $p < .001$ ; \*\* $p < .01$ ; \* $p < .05$

## Listing 1. Analysis R code

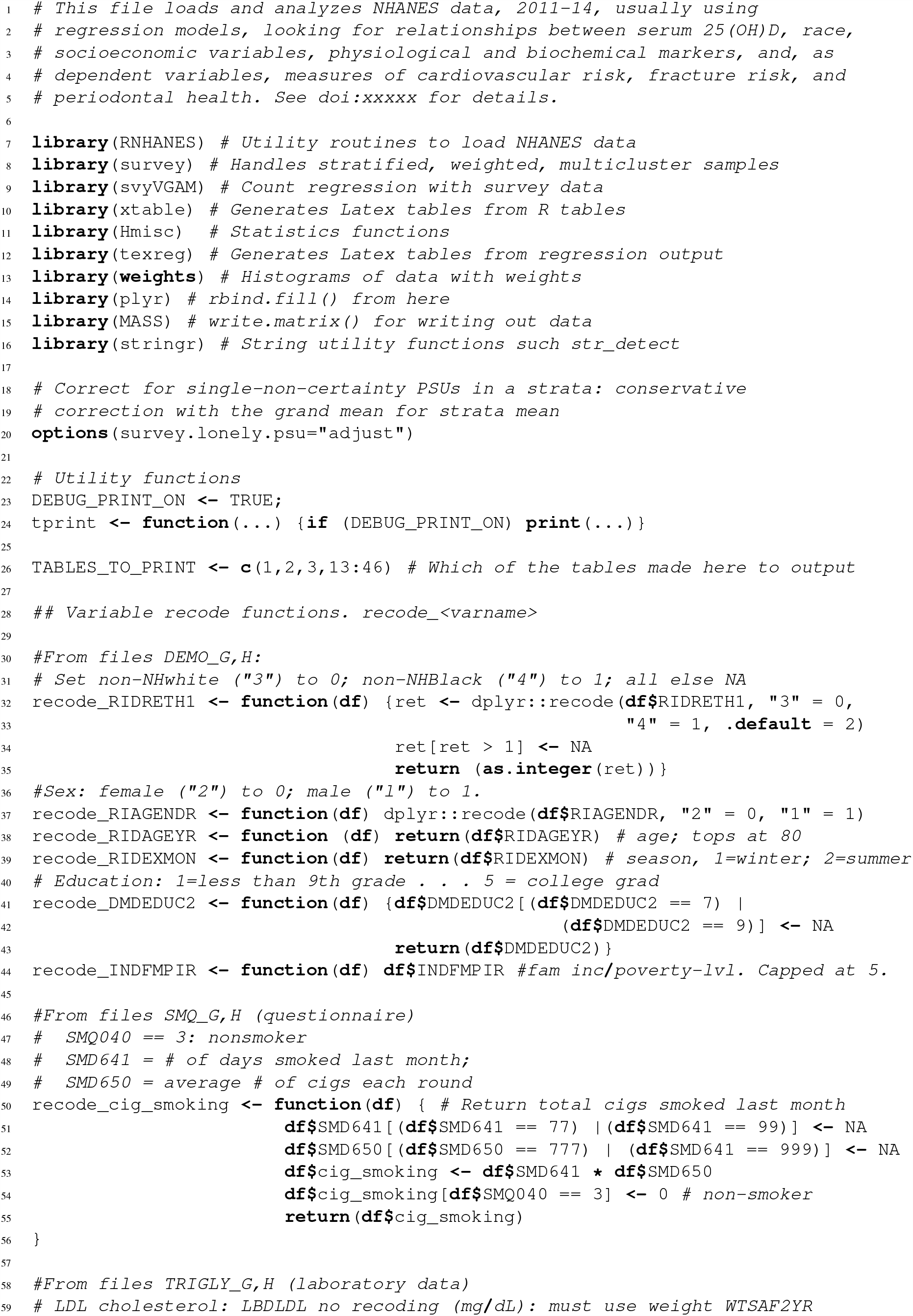

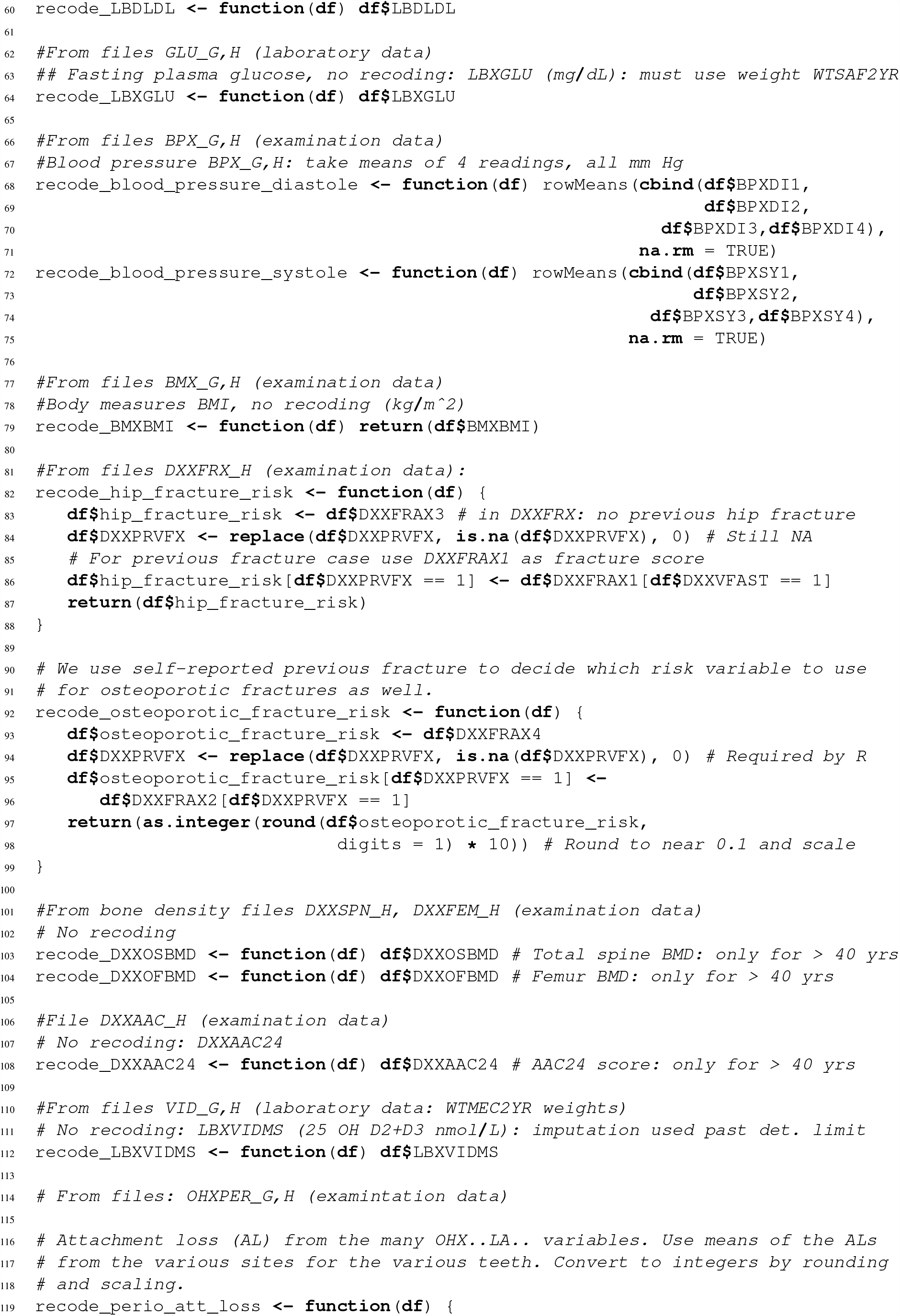

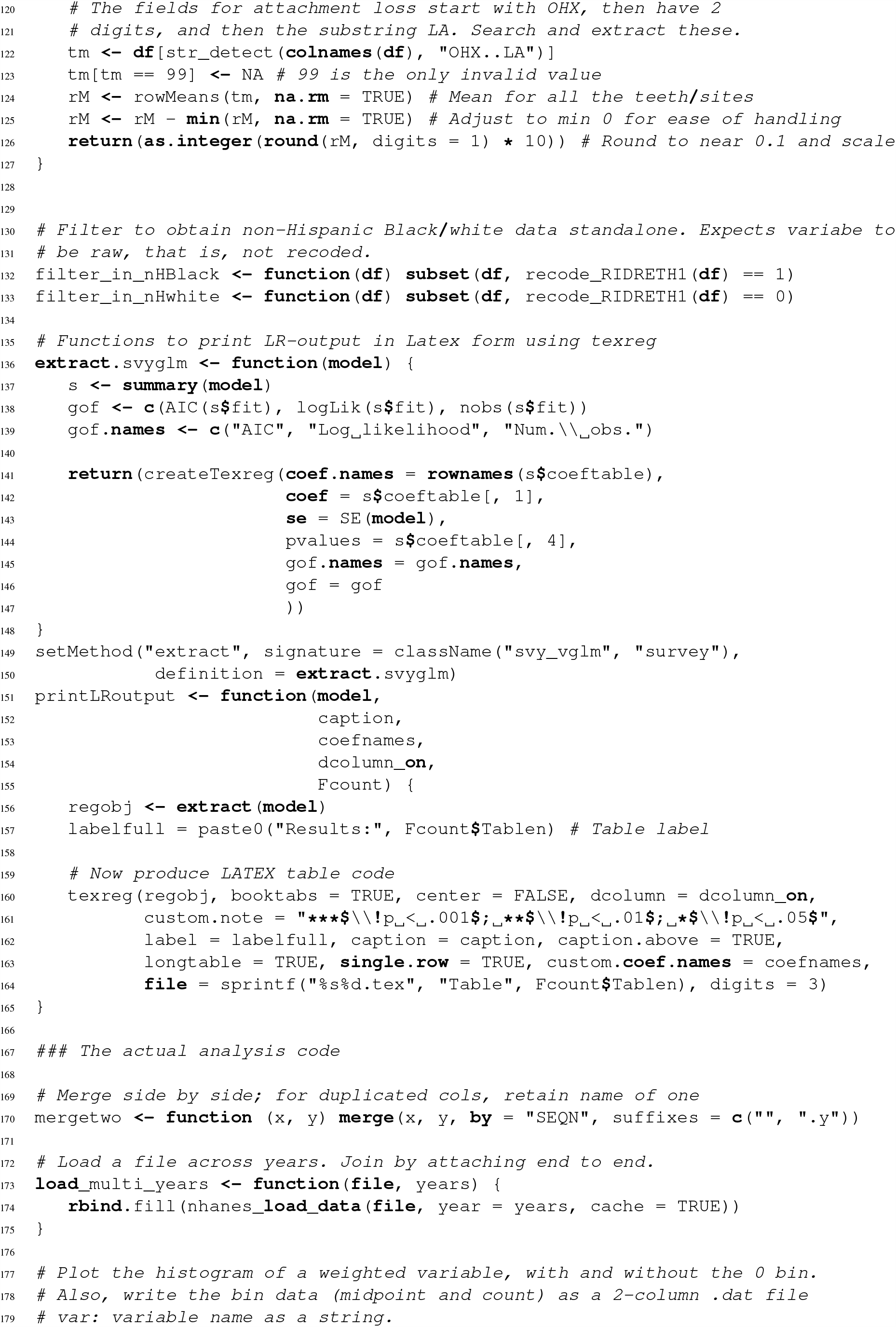

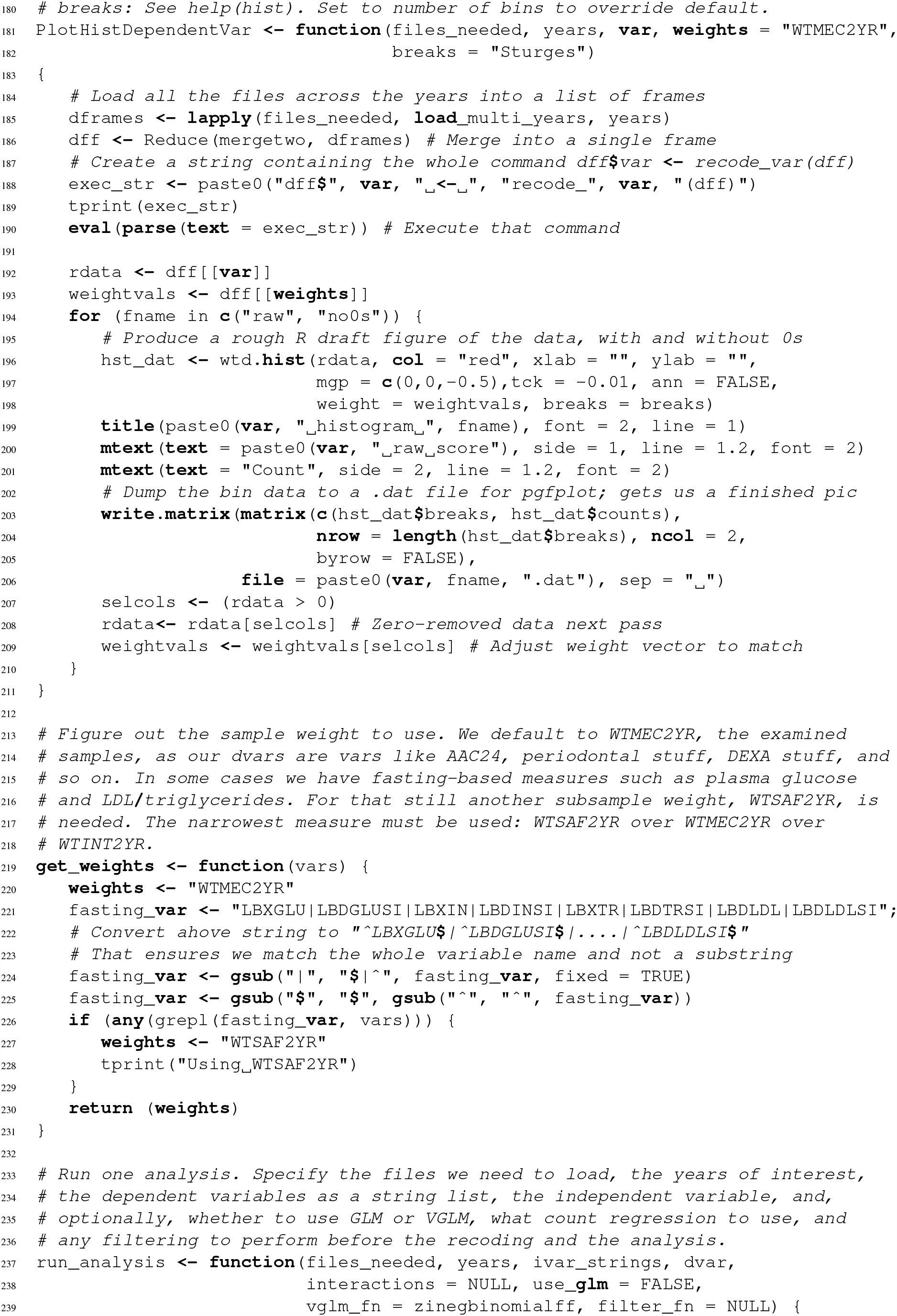

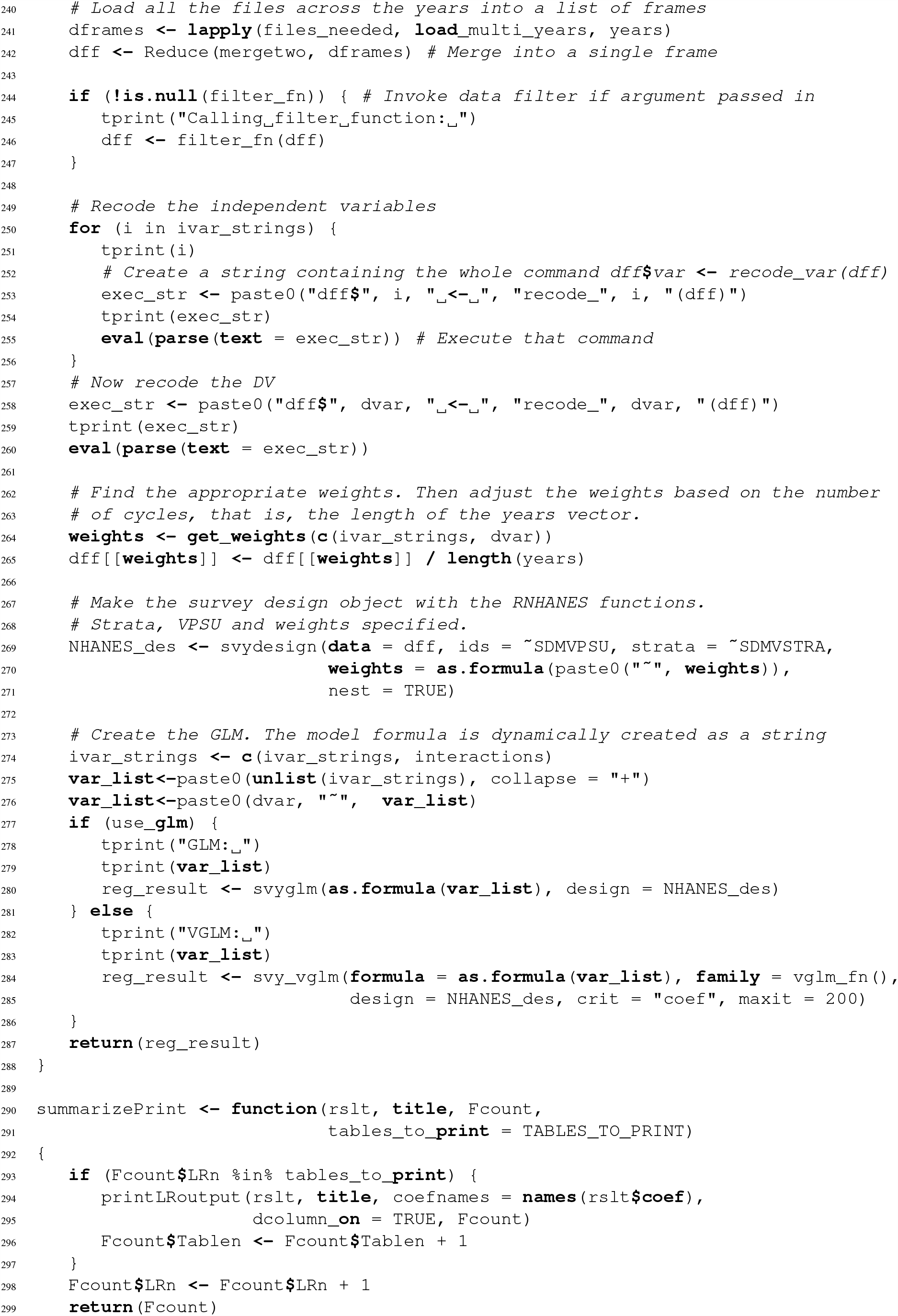

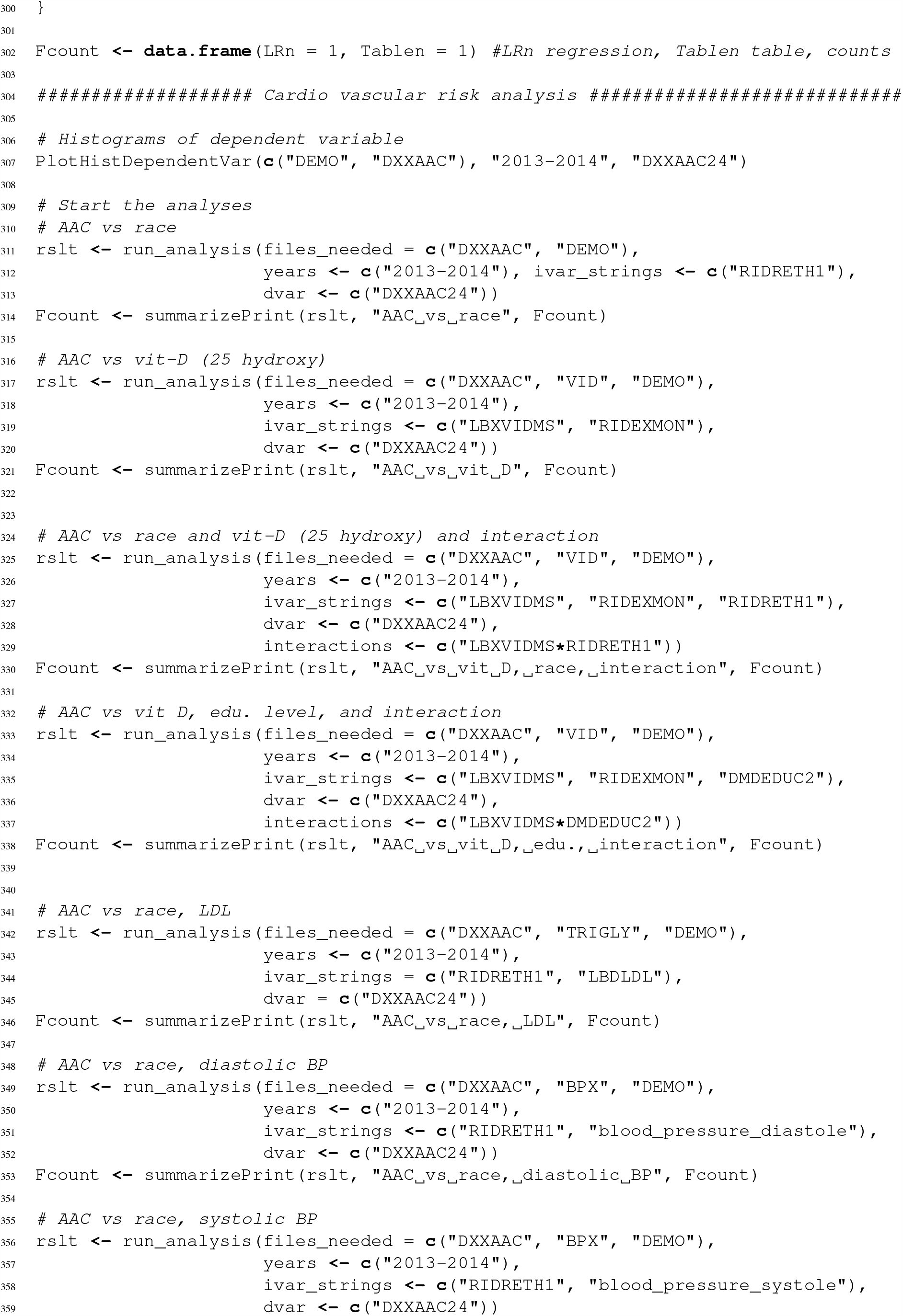

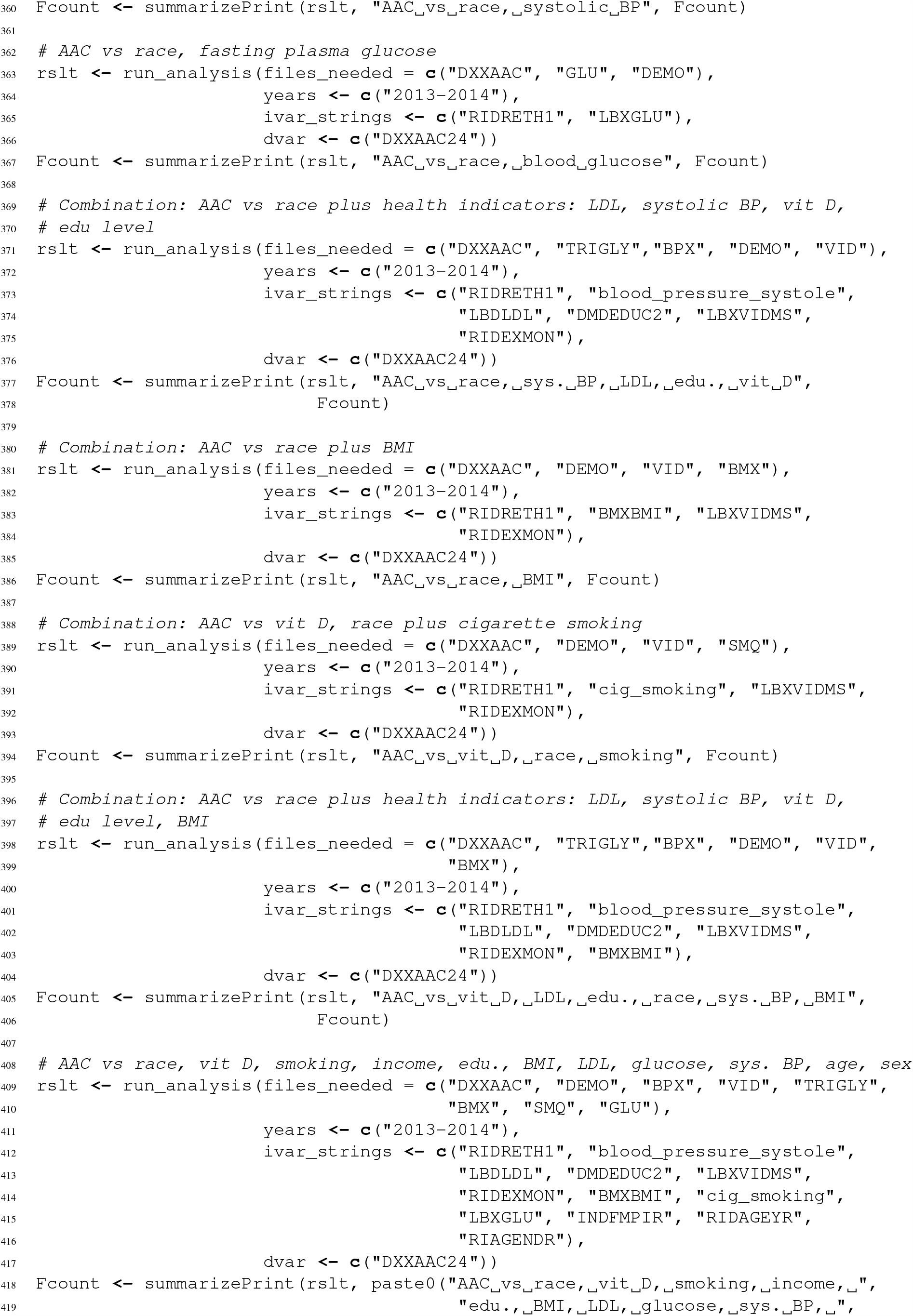

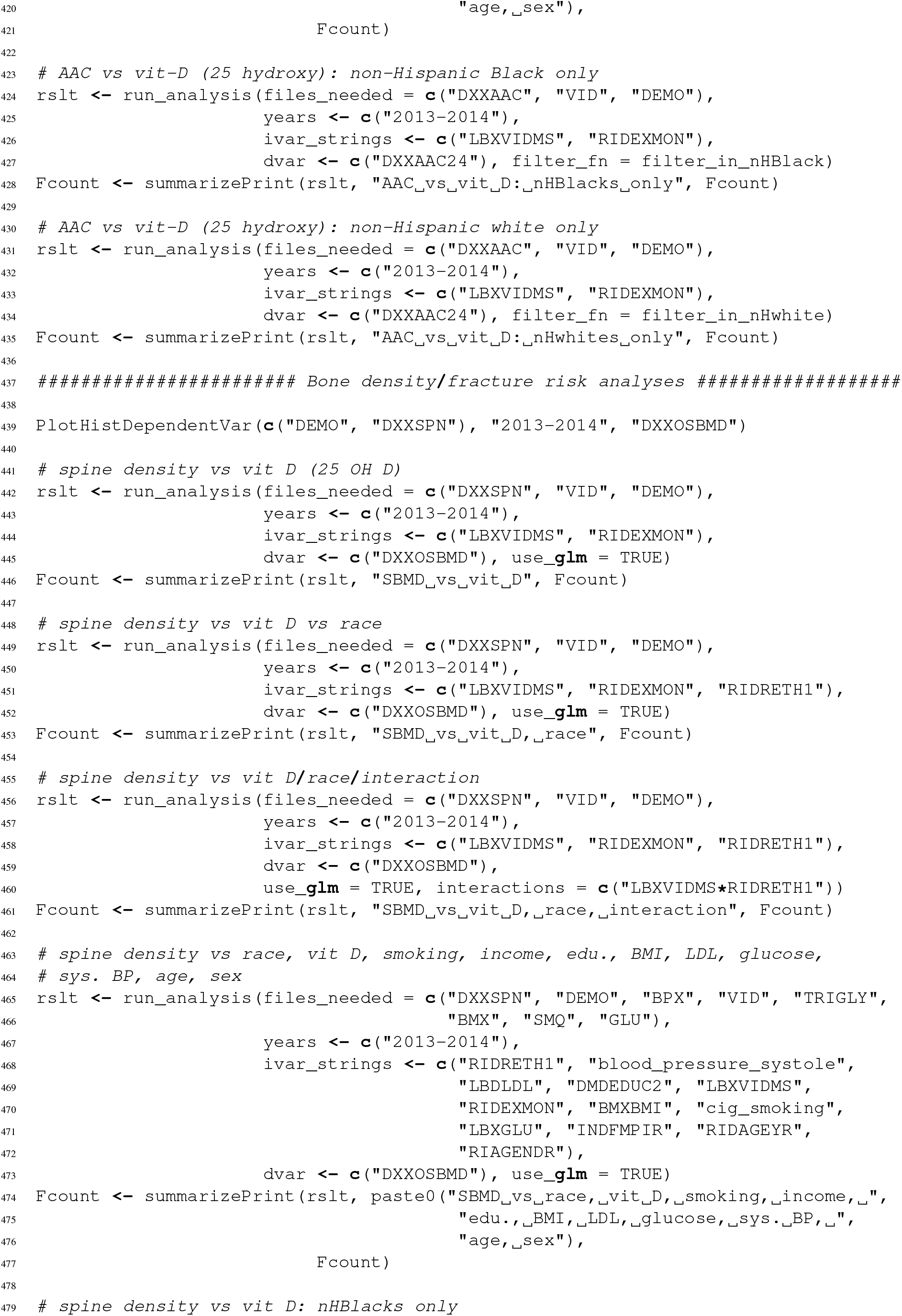

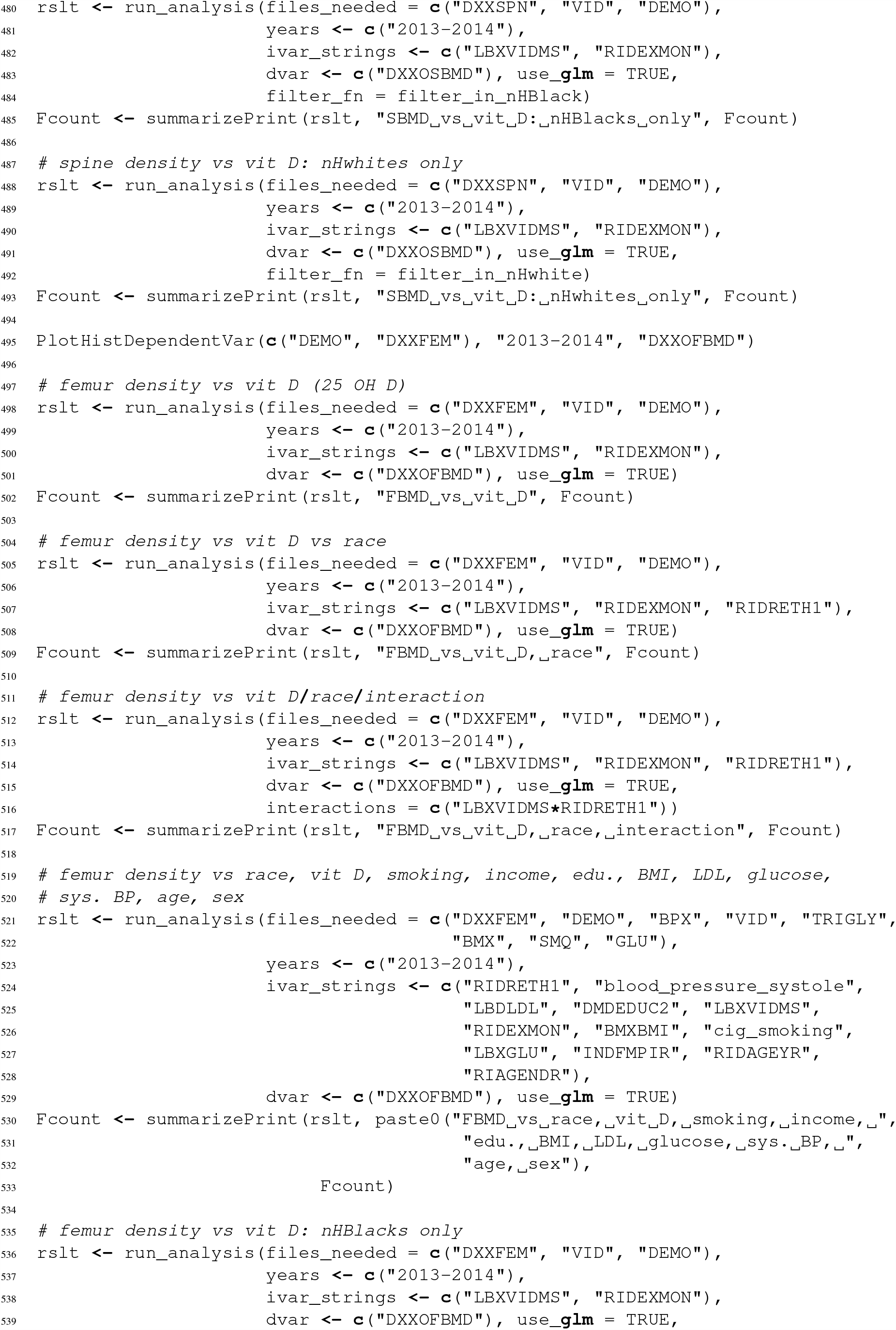

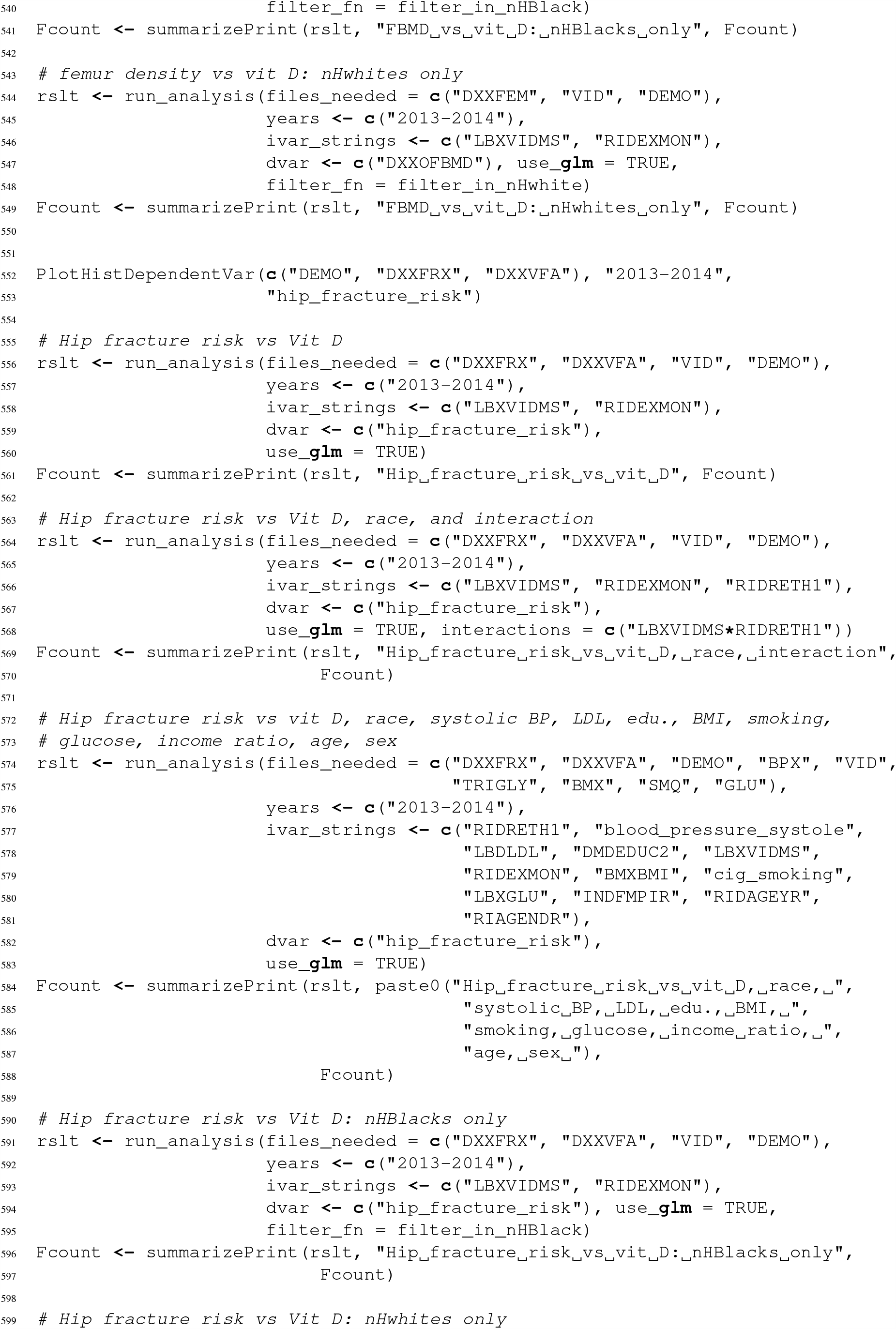

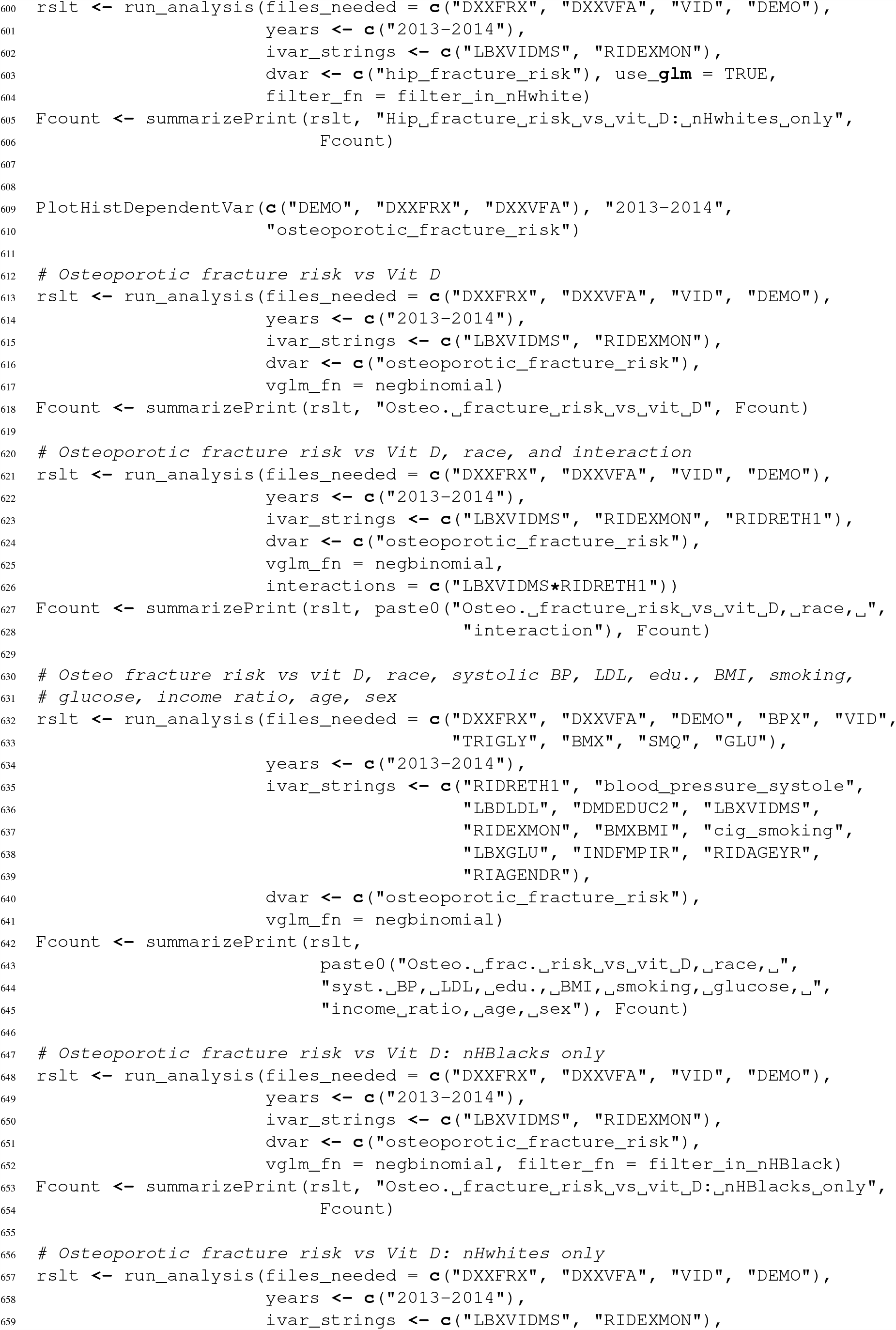

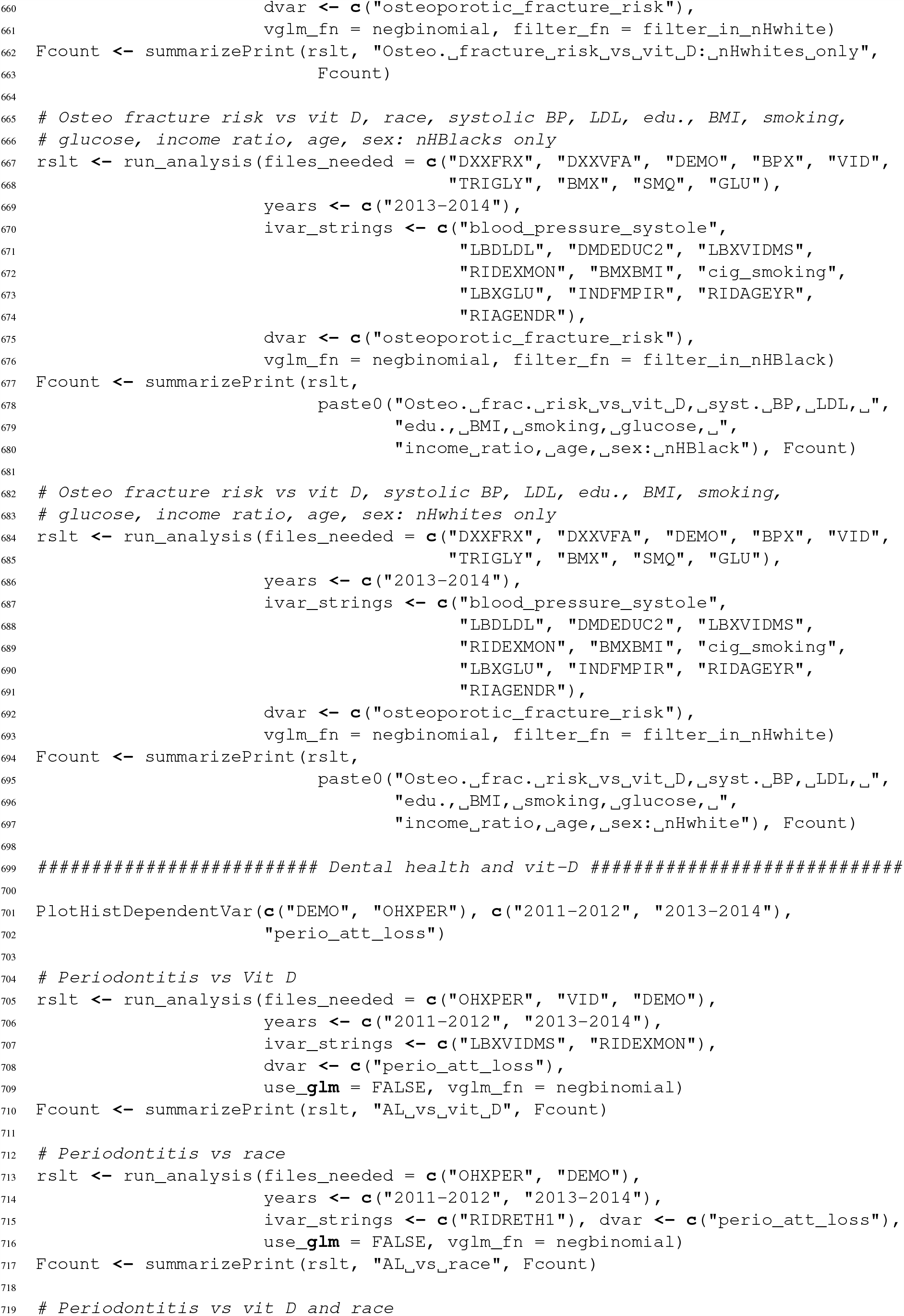

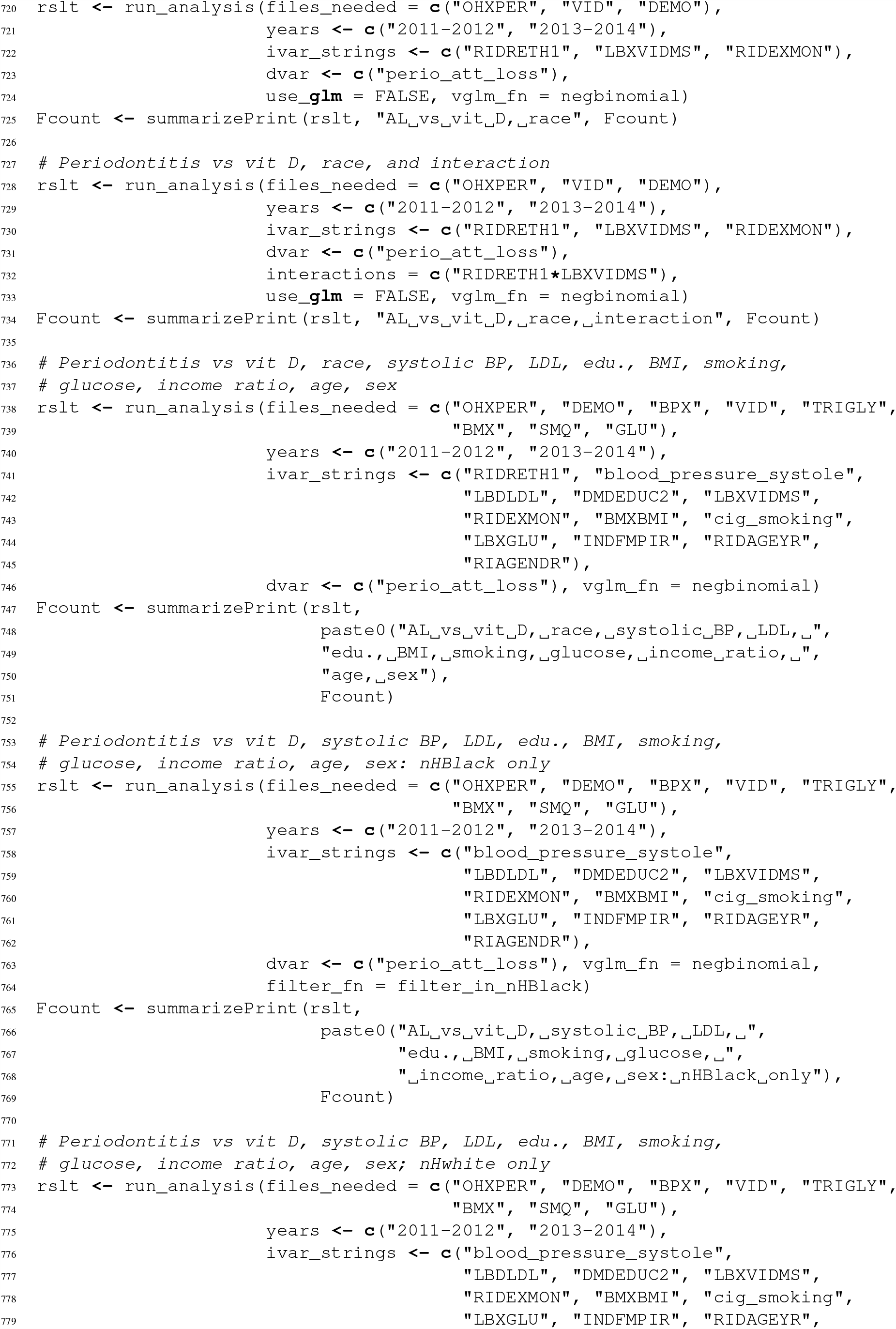

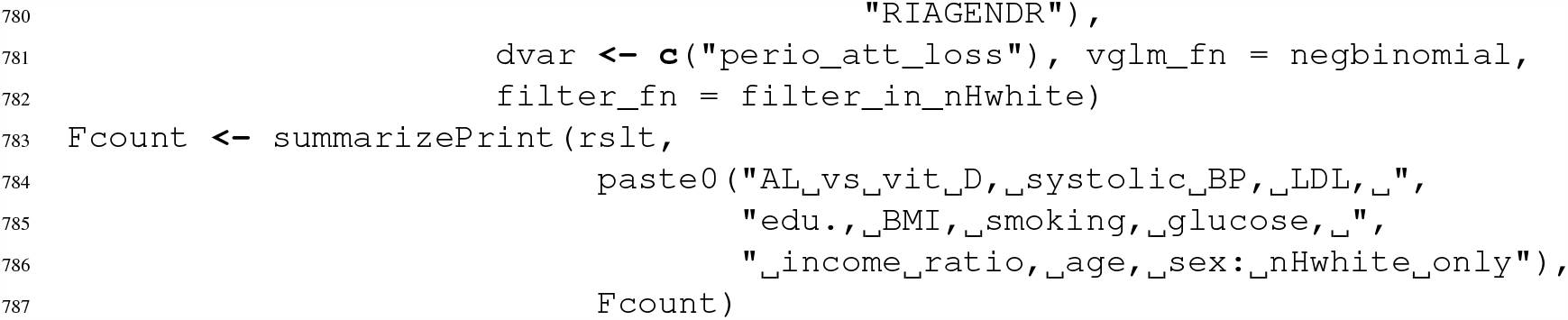

